# Impaired memory B-cell formation after mRNA-based COVID-19 booster vaccination in patients with inflammatory bowel disease receiving anti-TNF treatment

**DOI:** 10.64898/2026.08.28.26359302

**Authors:** Paul A. Gill, Lachlan R. Bradbury, Alina Wang, Julie Hogg, Kathryn Demase, Jo McKenzie, Holly A. Fryer, Daryl Geers, Luca M. Zaeck, Irene Boo, P. Mark Hogarth, Heidi E. Drummer, Rory D. de Vries, Robyn E. O’Hehir, Miles P. Sparrow, Menno C. van Zelm

**Author notes:** **Corresponding authors:** Paul A. Gill, PhD, Allergy and Clinical Immunology Laboratory, Monash University, 89 Commercial Road, Melbourne, Victoria 3004, Australia., Menno C. van Zelm, PhD, Department of Immunology, Erasmus University Medical Center, Dr. Molewaterplein 40, 3015 GD Rotterdam, the Netherlands.

## Abstract

**Background:** Patients receiving anti-TNF treatment for chronic inflammatory disease display impaired antibody responses, but it remains unclear how immune memory formation is affected. We evaluated antibody responses and memory B cells (Bmem) after COVID-19 booster vaccination in inflammatory bowel disease (IBD) patients receiving anti-TNF treatment.

**Methodology:** Blood was sampled at baseline, 1, and 6 months after WH1/BA.5 bivalent or XBB.1.5 monovalent vaccination from 27 IBD patients receiving intravenous anti-TNF and 44 controls. Neutralizing antibodies were measured using an infectious virus assay. SARS-CoV-2 spike receptor binding domain (RBD)-specific serum IgG was quantified by ELISA, and RBD-specific Bmem were immunophenotyped by flow cytometry using recombinant proteins from ancestral, Omicron BA.1, BA.5, XBB.1.5, and JN.1 variants.

**Results:** Serum IgG to vaccine RBD and neutralizing antibodies in patients increased pre to 1 month post-vaccination, but were lower than controls. Ancestral-, BA.5- and XBB.1.5-specific Bmem increased after vaccination but were significantly lower in patients than controls. Within RBD-specific Bmem, frequencies of recently activated CD21^lo^ cells were increased after vaccination, and were higher in patients than controls. Fewer antigen-specific Bmem in patients expressed IgG4, and more expressed IgG3 or IgD following vaccination. Following vaccination, more RBD-specific Bmem recognized multiple viral variants. However, patients had fewer Bmem that could bind to subvariants than controls.

**Conclusion:** Antibody and Bmem responses to COVID-19 booster vaccination in anti-TNF-treated IBD patients displayed reduced capacity, durability and cross-reactivity, suggesting impaired immune memory for protection against breakthrough infection. This supports the recommendation for annual booster vaccination to prevent severe disease and viral spread.

**Highlights:**

- IBD patients on anti-TNF biologics mount antibody responses and form vaccine-specific memory B cells following WH1/BA.5 bivalent or XBB.1.5 monovalent mRNA COVID-19 booster vaccination.
- Vaccine-specific antibody responses and memory B cell formation in patients are significantly lower than in controls at 1- and 6 months after vaccination, indicative of poorer peak response and durability.
- Despite multiple vaccinations, vaccine-specific memory B cells from patients display phenotypic alterations and reduced recognition of multiple SARS-CoV-2 Omicron subvariants, potentially a consequence of impaired germinal center responses in patients.

## INTRODUCTION

Coronavirus disease 2019 (COVID-19) remains an ongoing global health concern. Although mRNA- and adenoviral vector-based COVID-19 vaccines were initially very effective at preventing severe disease and hospitalization from SARS-CoV-2,^1,2^ subsequent Omicron sub-variants generated by the continued antigenic evolution of SARS-CoV-2 evade vaccine-elicited immune responses to cause breakthrough infections.^3^ Reduced public health and personal protective measures have resulted in continuous circulation of SARS-CoV-2 in the community.^4,5^ Therefore, vulnerable individuals may still be at risk from severe COVID-19 and possibly the development of long COVID.^6,7^ Furthermore, COVID-19 continues to impact hospital function and patient care due to the risk of nosocomial spread.^8^

People with inflammatory bowel disease (IBD), primarily Crohn’s disease and ulcerative colitis, may be at higher risk for severe COVID-19 due to their need for immunosuppressive treatment.^9^ Indeed, initial studies highlighted that IBD patients receiving anti-tumor necrosis factor (TNF), particularly in combination with immunomodulating drugs, displayed reduced antibody responses after SARS-CoV-2 infection.^10^ Furthermore, these IBD patients had impaired serological and cellular responses to primary COVID-19 vaccination,^11–13^ highlighting the important role of TNF-α in forming lymphoid structures crucial for robust humoral immune responses.^14^ Consequently, IBD patients treated with immunosuppressive therapies were prioritized to receive a third primary dose of the ancestral SARS-CoV-2 (Wuhan Hu-1; WH1) mRNA-based vaccine.^15^ Despite receiving an extra primary vaccine dose, antibody responses among IBD patients remained lower and less durable than those of healthy individuals, and IBD patients were found to have higher rates of breakthrough infections post-vaccination.^16, 17^ In addition to antibodies, memory B cells (Bmem) are critical for maintaining long-term immunity in the context of waning antibody responses.^18^ SARS-CoV-2 receptor-binding domain (RBD)-specific Bmem in healthy individuals are expanded after COVID-19 booster vaccination, and demonstrate greater durability than antibodies.^19,20^ Bmem induced by the original priming vaccination series using the ancestral COVID-19 vaccine typically display markers of class-switched Bmem (CD27^+^, IgD^-^) and predominantly express IgG1.^21,22^ Additional booster doses may increase the breadth of Bmem binding to SARS-CoV-2 Omicron variants.^23^ Taken together, the properties of vaccine-induced RBD-specific Bmem make these an important measure of the long-term immune response against severe COVID-19.^24,25^

The vaccine landscape for COVID-19 has continued to evolve with updated bivalent and subsequently monovalent mRNA-based booster vaccines modified to encode the spike RBD of emerging Omicron subvariants BA.1, BA.5, XBB.1.5 and JN.1.^26,27^ There were initial concerns that immune imprinting to the ancestral variant would impair the recognition of variants following updated COVID-19 booster vaccines.^28,29^ However, despite boosting ancestral specific responses, these vaccines also successfully directed the immune response towards the antigenically distinct variants, increasing protection against severe COVID-19.^23,27,30^ Still, limited evidence exists to demonstrate the immunocompetency of IBD patients treated with immunosuppressives to updated booster vaccine formulations. XBB.1.5 monovalent mRNA booster vaccination was found to increase neutralizing antibodies against JN.1 in 18 patients with IBD receiving anti-TNF treatment. However, it is unclear how this compares to the response in healthy individuals.^31^ In addition, there is limited information around the magnitude and quality of the RBD-specific Bmem response to booster vaccination in IBD patients receiving anti-TNF treatment. IBD patients on anti-TNF treatment have fewer spike-specific Bmem 3-6 months after double-dose mRNA-based primary vaccination than healthy individuals.^12^ Spike-specific Bmem frequencies remain lower than healthy individuals after a third dose booster, with reduced B-cell receptor (BCR) somatic hypermutation and IgG class-switched Bmem, suggesting reduced germinal center (GC) activity.^32^ However, previous studies have only examined Bmem responses to ancestral WH1 vaccines in small cohorts of IBD patients. It remains unclear if the Bmem response in IBD patients on anti-TNF treatment can also be directed to emerging variants after vaccination with updated boosters, and if phenotypic differences persist.

Taken together, there is a need for further studies to support evidence-based clinical recommendations for COVID-19 booster vaccination in patients receiving immunomodulating drugs combined with anti-TNF, particularly as recommendations vary around the world.^33–37^ Further clarity on vaccination guidelines is necessary to mitigate confusion in primary healthcare settings, especially as people in high-risk groups continue to experience anxiety about COVID-19.^38^

Here, we investigated the immunocompetency of 27 IBD patients receiving immunomodulating drugs combined with anti-TNF treatment following WH1/BA.5 bivalent or XBB.1.5 monovalent vaccination. Through analysis of antibody responses and Bmem formation after 1 and 6 months, we show that anti-TNF-treated IBD patients mounted vaccine responses, which were quantitatively lower than healthy individuals, highlighting the need for booster vaccinations to maintain protective immunity in this iatrogenic immunodeficient population.

## METHODS

### Study design

A longitudinal prospective study was conducted at the Alfred Hospital (Victoria, Australia) between March 2023 and January 2025 according to the Declaration of Helsinki and approved by local ethics committees (Alfred Health ethics 32/21, Monash University ethics 72794). Patients with IBD who were receiving intravenous anti-TNF (Infliximab) treatment were recruited after having made the decision to receive a COVID-19 booster vaccination. A total of 27 patients were included in the final analysis. Control samples were collected from healthcare workers at the Alfred Hospital (n=13) and as part of the SWITCH-ON study at Erasmus MC (n=31), as described before.^30^ The SWITCH-ON study protocol (MEC-2022-0462) was approved by the Medical Ethics Committee of Erasmus University Medical Center (Rotterdam, the Netherlands), the sponsor site, and the local review boards of other participating centers (Amsterdam University Medical Centers, the Leiden University Medical Center, and the University Medical Center Groningen). Participants were sampled at baseline, and 1 month and 6 months after booster vaccination with either a WH1/BA.5 bivalent or XBB.1.5 monovalent mRNA-based COVID-19 mRNA vaccine. Basic demographics including age, sex, COVID-19 vaccination and SARS-CoV-2 infection history were recorded. Patient clinical and treatment information was also collected during the study.

### Blood sampling and processing

Participants underwent venipuncture for collection of up to 40 mL of blood at each sampling timepoint. Blood samples were processed as described previously.^39,40^ Briefly, a 200 µL aliquot of whole blood was used for whole blood cell count using a Cell-dyn analyzer (Abbott Core Laboratory, Abbott Park, IL) and Trucount analysis. The remaining blood sample was separated to isolate and store plasma (Alfred hospital samples) and serum (Erasmus MC samples) at –80 °C. Peripheral blood mononuclear cells (PBMC) were isolated by Ficoll-Paque density gradient centrifugation and cryopreserved in liquid nitrogen for later analysis.

### Measurement of SARS-CoV-2 neutralizing antibodies

Neutralizing antibodies (NAb) in plasma and serum samples from patients and controls were measured to ancestral, Omicron BA.5, XBB.1.5, and JN.1 using an infectious virus assay as previously described.^41,42^ The following SARS-CoV-2 viral isolates were used: WH1, hCoV-19/Australia/NSW2715/2020; BA.5, hCoV-19/Australia/NSW-ICPMR-29513/2022; XBB.1.5, hCoV-19/Australia/NSW-ICPMR-44477/2023; JN.1, hCoV-19/Australia/NSW-ICPMR-54023/2024. Briefly, clonal HAT-24 cells were generated by transducing lentiviral particles into HEK293T (R70007, Thermo Fisher, Waltham, MA) cells to stably express ACE2 and TMPRSS2 receptors. Heat-inactivated test samples were half-log serially diluted with DMEM-5% FBS. An equal volume of SARS-CoV-2 virus, at twice the median lethal dose, was added to the diluted sample. Sample-virus mixture was incubated for 1 hr at 37 °C and then added to plated Hoechst-33342 stained HAT-24 cells in 384 well plates. Plates were incubated for 20 hours at 37 °C, 5% CO_2_ before enumerating nuclear counts with high-content fluorescence microscopy (ImageXpress Pico Cell Imaging System, Molecular Devices, San Jose, CA) and CellReporterXpress software. Neutralization was expressed as the reciprocal of the dilution of plasma or serum required to inhibit 50% viral entry (ID50), calculated from the nonlinear regression line. The lowest neutralization titer detected by the assay was 20, with neutralization titers <20 arbitrarily expressed as 10.

### Protein production and tetramerization

Recombinant SARS-CoV-2 nucleocapsid (NCP) and spike RBD proteins from ancestral WH1, Omicron BA.1, BA.5, XBB.1.5 and JN.1 was generated as previously described.^30,39^ Briefly, DNA constructs containing an N-terminal Fel d 1 leader sequence, C-terminal AviTag and 6-His affinity Tag were cloned into a pCR3 plasmid and produced using the Expi293 expression system (Thermo Fisher) and purified using cobalt affinity column purification. Purified proteins were used for ELISAs. Aliquots of purified RBD were biotinylated and tetramerized with fluorochrome-conjugated streptavidin. The following fluorescent tetramers were used in analysis of all samples: [RBD WH1]_4_-BUV395, [RBD WH1]_4_-BV421, [RBD BA.1]_4_-BUV496 and [RBD JN.1]_4_-BV650. Recipients of WH1/BA.5 bivalent vaccine were also stained with [RBD BA.5]_4_-BUV737, [RBD BA.5]_4_-BUV480 and [RBD XBB.1.5]_4_-BUV615, whilst XBB.1.5 monovalent recipients used [RBD XBB.1.5]4-BUV737, [RBD XBB.1.5]4-BUV480 and [RBD BA.5]4-BUV615 (**Supplementary Tables 1 and 2**).

### Detection of SARS-CoV-2-specific IgG antibodies using ELISA

Quantitation of IgG specific for ancestral NCP and WH1, Omicron BA.1, BA.5, XBB.1.5 and JN.1 RBD was performed using an in-house ELISA protocol previously described.^19^ Ninety-six well EIA-RIA plates (Corning Incorporated, Costar, St Louis, MO) were coated with 2 μg/mL recombinant unbiotinylated protein monomers overnight at 4°C. To allow quantitation for IgG concentration, a standard curve was prepared by serially diluting recombinant human IgG (in-house made human Rituximab). Plates were blocked with 3% BSA in PBS and subsequently incubated with plasma or serum samples serially diluted from 1:30 to 1:10,000 for quantitation of RBD-specific IgG, and 1:30 for NCP-specific IgG. Antigen-specific IgG was detected using rabbit anti-human IgG HRP (Dako, Glostrup, Denmark). ELISA plates were developed using TMB solution (Life Technologies, Carlsbad, CA), and the reaction was stopped with 1M HCl. Absorbance (OD450nm) was measured using a Multiskan Microplate Spectrophotometer (Thermo Fisher). RBD-specific IgG concentrations were interpolated from the standard curve using GraphPad Prism (v10, Dotmatics, Boston, MA). Titration curves were also generated, and the area under the curve (AUC) was calculated for each variant calculated using GraphPad Prism. Total variant binding capacity was determined by summation of all variant AUC values.

### Assessment of absolute numbers of leukocyte and lymphocyte subsets

Absolute numbers of major leukocyte and lymphocyte subsets were assessed using a lyse-no-wash method as previously described.^19^ In short, 50 µL of whole blood was incubated with an antibody cocktail (**Supplementary Table 1**) in a BD Trucount tube (BD Biosciences) prior to addition of 1x BD lysis solution (BD Biosciences). Samples were then acquired on a FACSLyric analyzer (BD Biosciences) and data analyzed using FlowJo^TM^ software (v10.10.0, BD Biosciences). Trucount analysis was conducted on all IBD patient samples and a subset of control samples (**Supplementary Table 1**). Absolute counts of B cells were used to calculate absolute numbers of RBD-specific Bmem.

### Immunophenotyping of memory B cells using flow cytometry

Fluorescent streptavidin-RBD protein tetramers were incorporated into a 20-color spectral flow panel cytometry panel for Bmem phenotyping (**Supplementary Tables 1 and 2**). Five to fifteen million thawed PBMC were incubated at room temperature in the dark for 15 minutes in a total volume of 250 μL with PBS containing 0.1% LIVE/DEAD^TM^ Fixable blue stain (Thermo Fisher). After washing in FACS buffer (PBS with 0.1% sodium azide and 0.2% BSA), PBMC were stained in 250 μL FACS buffer with the following antibodies: CD3, CD11c, C19, CD21, CD27, CD38, CD71, IgM, IgD, IgG1, IgG2, IgG3, IgG4, IgA, and 5 μg/mL of each in-house generated RBD fluorescent tetramer (**Supplementary Table 2**). In a separate tube, 1–5 million PBMC were also stained in 100 µL PBS with 0.1% LIVE/DEAD^TM^ Fixable blue stain, washed and then incubated in a total volume of 100 μL with FACS buffer containing CD3, CD19, CD27, IgD and fluorochrome-conjugated streptavidin (BUV395, BV421, BV480, BUV737, BUV496, BUV615 and BV650) without respective RBD proteins (**Supplementary Table 2**). Following staining, cells were washed, fixed with 2% PFA for 20 min at room temperature in the dark, and washed once before acquisition on a 5-laser Cytek Aurora (Cytek Biosciences, Fremont, CA) using SpectroFlo® software (v3.3.0, Cytek Biosciences). Data analysis was performed using FlowJo™ Software (v10.10.0).

### Data analysis and statistics

Statistical analysis was performed using GraphPad Prism (v10). The non-parametric Wilcoxon matched-pairs signed-rank test was used for paired data, the non-parametric Mann-Whitney U test for unpaired data, and the Chi-squared test for categorical data. Gaussian generalized linear mixed modelling (GLMM) analysis was performed using R (v4.5.2) within RStudio (v2025.09.2, Build 418) to assess the impact of sex, primary vaccination regime, age, NCP seropositivity and vaccination dose on Yeo-Johnson transformed data for RBD-binding antibodies, neutralizing antibodies, RBD-specific antibody variant binding capacity, RBD-specific Bmem frequency and RBD-specific Bmem variant binding capacity in IBD patients and healthy controls. Additional GLMM analysis was performed to assess the impact of clinical diagnosis, treatment regime and infliximab level on IBD patient RBD-specific antibodies and RBD-specific Bmem. Radar plots and analysis were generated using the fmsb package (v0.7.6) within RStudio. For all tests, *p* < 0.05 was considered significant.

## RESULTS

### Cohort characteristics

A total of 27 IBD patients being treated with intravenous anti-TNF (median age 38, 41% female), and 44 healthcare workers (median age 51, 70% female) were included (**Table 1**). The IBD cohort included significantly fewer females (41%) than the control cohort (70%; p<0.05), and the median age was significantly lower than the healthy cohort. The IBD cohort consisted of 18 individuals diagnosed with Crohn’s disease and 9 individuals diagnosed with ulcerative colitis. The treatment regime for intravenous infliximab varied between 5 mg/kg every 8 weeks and 10 mg/kg every 4 weeks, with 56% of the patients receiving an additional immunomodulating drug (**Supplementary Table 3**).

**Table 1.** Cohort Characteristics.

| Cohort |  | IBD | Control |
| --- | --- | --- | --- |
| Sample Size |  | 27 | 44 |
| Female (%) |  | 11 (41%) | 31 (70%)* |
| Median Age at recruitment (range) |  | 38 (22-77) | 51 (22-65)** |
| IBD Diagnosis | Crohn's disease | 18 (66.7%) | N/A |
|  | Ulcerative Colitis | 9 (33.3%) | N/A |
| Study Booster Vaccine | Bivalent BA.1 | 1 (4%) | 0 (0%) |
|  | Bivalent BA.4/5 | 19 (70%) | 22 (50%) |
|  | Monovalent XBB.1.5 | 7 (26%) | 22 (50%) |
| Study Vaccine Number | Three | 2 (7%) | 2 (5%) |
|  | Four | 15 (56%) | 18 (41%) |
|  | Five | 8 (30%) | 20 (45%) |
|  | Six | 0 (0%) | 4 (9%) |
|  | Seven | 2 (7%) | 0 (0%) |
| Formulation of Primary Vaccination | mRNA | 21 (78%) | 18 (41%)** |
|  | Adenoviral | 6 (22%) | 26 (59%) |
| Median duration in days since last vaccine (range) |  | 432 (196-828) | 350 (173-744)**** |
| Median duration in days between vaccination and 1M post sample (range) |  | 30 (16-70) | 28 (23-53) |
| Median duration in days between vaccination and 6M post sample (range) |  | 203 (92-244) | 176 (138-259) |
| Nucleocapsid seropositive at baseline (%) |  | 6 (22%) | 17 (39%) |
| Nucleocapsid seropositive after vaccination (%) |  | 2 (7%) | 12 (27%)* |

Statistical significance as determined by Mann-Whitney U test for continuous data, Chi-squared test for categorical data. \*P<0.05, \*\*P<0.01, \*\*\*\*P<0.0001

One IBD patient received the Omicron WH1/BA.1 bivalent vaccine, 19 the WH1/BA.5 bivalent vaccine, and 7 the XBB.1.5 monovalent vaccine. In the control group, 22 received the WH1/BA.5 bivalent, and 22 the XBB.1.5 monovalent vaccine. For the majority of IBD patients, this was their fourth (56%) or fifth (30%) COVID-19 vaccine dose. This was similar to the controls with 41% receiving dose 4 and 45% dose 5. A larger proportion of IBD patients received an mRNA-based vaccine as part of their primary double-dose vaccination regime than the controls (78% vs 41%, p<0.01). Controls had a shorter median duration since their previous vaccine dose than the IBD patients (350 vs 471 days, p<0.0001).

Peripheral blood was collected from patients and controls at baseline, and 1 month (IBD patients: median 30 days; controls: median 28 days) and 6 months (IBD patients: median 203 days; controls: median 176 days) after COVID-19 booster vaccination (**Table 1** and **Figure 1A**). One IBD patient who received the XBB.1.5 monovalent vaccine did not have peripheral blood collected 6 months after vaccination but still had the baseline and 1 month post-vaccination sample included in the final analysis. At baseline, 39% of controls and 22% of IBD patients were seropositive for SARS-CoV-2 NCP. Furthermore, an additional 27% of controls and 7% of IBD patients were newly NCP seropositive at sampling 1 month after vaccination, suggesting potential infection during the study (**Table 1** and **Supplementary Figure 1A**).

**Figure 1.**
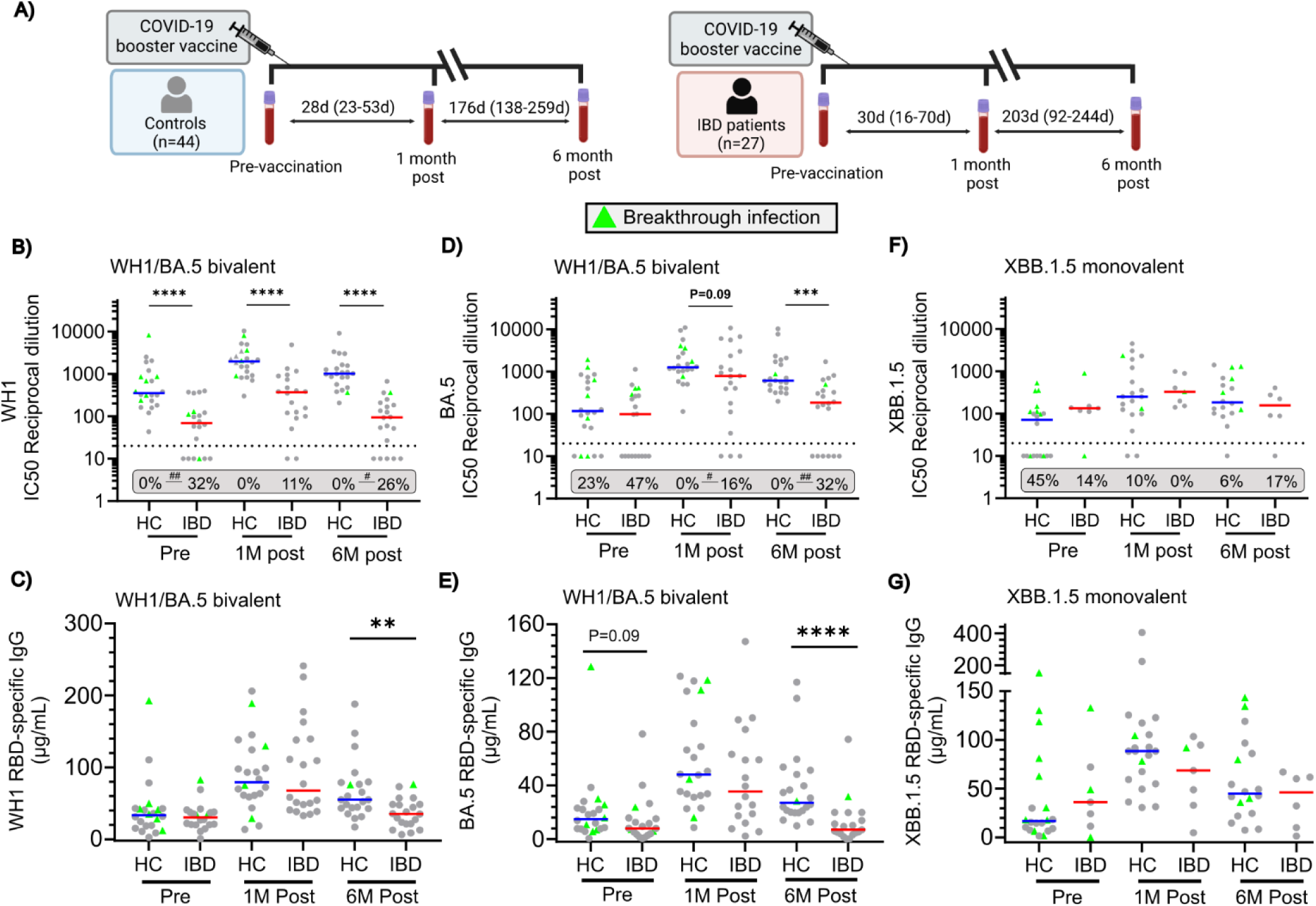
Antibody response to WH1/BA.5 bivalent or XBB.1.5 monovalent COVID-19 vaccination in IBD patients and controls. **(A)** Schematic of study design with sampling timepoints. **(B)** Neutralizing antibody titers against ancestral (WH1) SARS-CoV-2, **(C)** WH1 RBD-specific IgG, **(D)** neutralizing antibody titers against Omicron BA.5, and, **(E)** BA.5 RBD-specific IgG concentrations, in WH1/BA.5 bivalent vaccinated IBD patients (n=19) and controls (HC)(n=22). **(F)** Neutralizing antibody titers against XBB.1.5, and, **(G)** XBB.1.5 RBD-specific IgG concentrations in XBB.1.5 monovalent vaccinated IBD patients (n=7) and HCs (n=22). Numbers in grey box represent proportions of individuals below cutoff value of 20 (dotted line) without neutralizing antibodies. Statistical significance between proportions of non-neutralising responses between patients and controls as calculated by Chi-squared test. #p<0.05, ##p<0.01. Horizontal lines depict medians. Breakthrough infections were determined as seropositivity to SARS-CoV-2 Nucleoprotein (NCP). Statistical significance between patients and controls as calculated by Mann-Whitney test. *p<0.05, **p<0.01, ***p<0.001, ****p<0.0001.

### Reduced antibody response in IBD patients after COVID-19 booster vaccination

We first assessed serum antibody responses to vaccine antigens by measuring plasma NAb and RBD-binding IgG levels. IBD patients that received the WH1/BA.5 bivalent vaccine had a significant increase (69.3 vs 367.1 IC50, p=0.002) in NAb titers against WH1; however, NAb titers declined over time and were significantly lower (367.1 vs 94.5 IC50, p=0.0003) at 6 months post-vaccination (**Figure 1B**). Furthermore, NAb titers against WH1 were significantly lower in IBD patients than controls, with a significantly greater proportion of patients without NAb titers against WH1. WH1/BA.5 bivalent vaccination significantly increased WH1 RBD-specific IgG levels (30.6 vs 67.8 µg/mL, p<0.0001). The durability of WH1 RBD-specific IgG was poorer in patients receiving WH1/BA.5 bivalent vaccination, with significantly lower WH1 RBD-specific IgG at 6 months post-vaccination due to a larger decline in antibody levels from 1 month to 6 months post-vaccination (**Figure 1C**, **Supplementary Figure 1B**). Similarly, NAb titers against BA.5 increased (97.7 vs 780.7 IC50, p<0.0001) in IBD patients that received the WH1/BA.5 bivalent vaccine (**Figure 1D**). Despite similar NAb titers against BA.5 before vaccination, IBD patients had lower NAb titers against BA.5 than controls after vaccination, with a significantly greater proportion of patients without NAb titers against BA.5 both 1 month and 6 months post-vaccination. BA.5 RBD-binding IgG was also significantly increased (7.9 vs 35.4 µg/mL, p=0.0003) in IBD patients who received the WH1/BA.5 bivalent vaccine (**Figure 1E**). IBD patients had significantly lower BA.5 RBD-binding IgG 6 months post-vaccination than controls, and a larger decease from 1 month to 6 months post-vaccination (**Supplementary Figure 1B**).

IBD patients who received the XBB.1.5 monovalent vaccine had significantly higher XBB.1.5NAb titers (p=0.03) and but not RBD-specific IgG levels (p=0.22) after 1 month (**Figure 1F,G**). Both NAb titers (p=0.05) and RBD-binding IgG antibody levels (p=0.03) decreased significantly between 1 month and 6 months post-vaccination, as was observed for controls (**Figure 1F,G**)(**Supplementary Figure 1C**).

Due to differences in demographics, infection and vaccination history between the IBD patients and controls, we performed additional GLMM analysis. Baseline RBD-specific IgG levels were found to be significantly higher in individuals who were seropositive for SARS-CoV-2 NCP prior to vaccination (β: 0.64, p=0.04). Furthermore, those who received a primary vaccination with an mRNA-based vaccine had significantly higher RBD-binding IgG to the primary antigen contained in the vaccine (i.e. BA.5 RBD in BA.5 bivalent and XBB.1.5 RBD in XBB.1.5 monovalent) at 1 month post-vaccination (β: 1.48, p=0.04). The number of previous vaccines received had a significantly positive impact on primary antigen RBD-binding IgG levels at 6 months post-vaccination (β: 0.69, p=0.048), which may have contributed to the improved durability in the antibody response observed in patients who received the XBB.1.5 monovalent vaccine as compared to the bivalent WH1/BA.5 vaccine. No specific IBD patient factors (disease type, concomitant immunomodulator treatment or Infliximab titer levels) were found to significantly impact changes to RBD-specific IgG levels. Whilst these additional factors influenced antibody responses, these were still significantly lower in IBDs than in controls. Despite IBD patients mounting antibody responses to booster vaccination, these responses were poorer in magnitude and durability than in controls.

### IBD patients have fewer RBD-specific Bmem

Spectral flow cytometry was conducted to evaluate circulating B cells before and after COVID-19 booster vaccination (**Supplementary Figure 2**). Total B-cell numbers did not change over time and were not different between IBD patients and controls (**Supplementary Figure 3A**). Transitional, naive mature B cells, and Bmem were not different between IBD patients and controls over time, except for significantly higher Bmem at 6 months post-vaccination in controls (**Supplementary Figure 3B-D**). Absolute numbers of plasmablasts were significantly higher in IBD patients than in controls across all timepoints **(Supplementary Figure 3E).** We next assessed if vaccination altered the activation profile of total Bmem in IBD patients and controls. Upregulation of CD71 is indicative of recent activation, whilst a reduction in CD21 expression occurs upon antigen recognition.^43,44^ Expression of CD71 was similar on total Bmem; however, IBD patients had increased frequencies of CD21^lo^ Bmem across all timepoints, indicating that this phenotype was likely influenced by their underlying disease (**Supplementary Figure 4A,B**).^45^ IBD patients also displayed altered maturation of IgG^+^ Bmem, with higher frequencies of IgG^+^ Bmem expressing CD27 before and 1 month post-vaccination (**Supplementary Figure 4C**). Furthermore, quantification of Ig isotypes and IgG subclasses of total Bmem highlighted that patients had significantly higher frequencies of IgG2^+^ and IgG3^+^ Bmem, and lower frequencies of IgG4^+^ Bmem (**Supplementary Figure 4D**). To examine vaccine-specific Bmem formation following COVID-19 booster vaccination, double-discrimination gating was used to identify RBD-specific cells within total Bmem **(Figure 2A)**. For recipients of the WH1/BA.5 bivalent vaccine, three populations of RBD-specific Bmem were defined: WH1^+^ only (WH1^+^BA.5^-^), WH1^+^BA.5^+^ cross-reactive, and BA.5^+^ only (WH1^-^BA.5^+^) (**Figure 2A**). In IBD patients and controls, the proportions of cross-reactive WH1^+^BA.5^+^ (IBD: 0.019 vs 0.070 % overall Bmem, p=0.0008; control: 0.067 vs 0.146 % overall Bmem, p=0.0001) and BA.5^+^ only RBD-specific Bmem (IBD: 0.024 vs 0.067 % overall Bmem, p=0.002; control: 0.164 vs 0.266 % overall Bmem, p<0.0001) were significantly increased 1 month after vaccination, followed by a significant decline from 1 month to 6 months post-vaccination (**Figure 2B**). Absolute numbers of cross-reactive and BA.5^+^ only RBD-specific Bmem also increased in IBD patients who received the BA.5 bivalent vaccine (**Supplementary Figure 5A**). However, the proportions of all three populations of RBD-specific Bmem in IBD patients were significantly lower pre-vaccination, and at 1 and 6 months post-vaccination, than in controls (**Figure 2B**).

**Figure 2.**
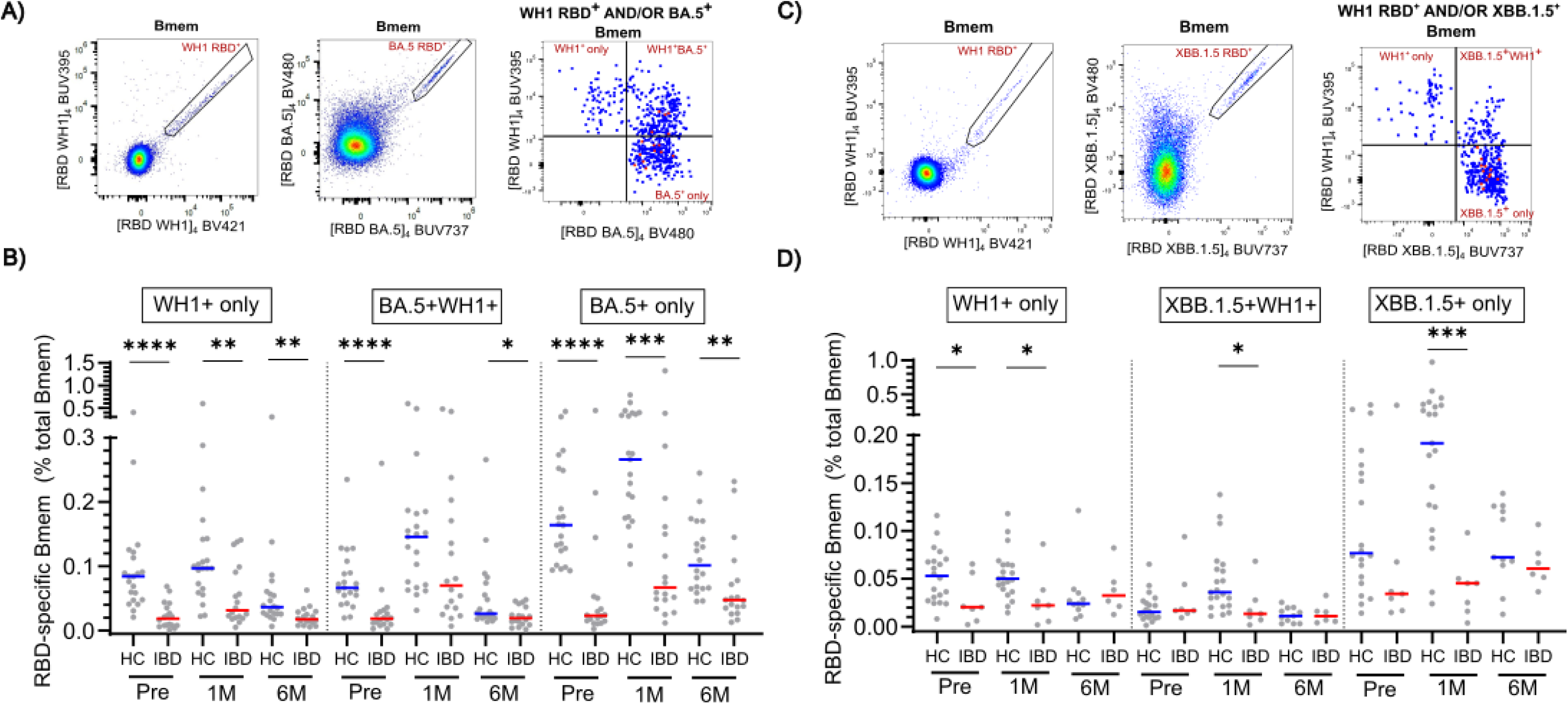
RBD-specific Bmem pre- and post COVID-19 booster vaccination in IBD patients and controls. **(A)** Gating strategy to determine WH1 and BA.5 specific Bmem in BA.5 vaccinated IBD patients (n=19) and controls (HC)(n=22), **(B)** WH1+ only, WH1+BA.5+ and BA.5+ only Bmem as a proportions of total Bmem before and after BA.5 bivalent vaccination, **(C)** gating strategy to determine WH1 and XBB.1.5 specific Bmem in XBB.1.5 monovalent vaccinated IBD patients (n=7) and HCs (n=22), **(D)** WH1+ only, WH1+XBB.1.5+ and XBB.1.5+ only Bmem as proportions of total Bmem before and after XBB.1.5 monovalent vaccination. Horizontal lines depict medians. Statistical significance between patients and controls as calculated by Mann-Whitney test. *p<0.05, **p<0.01, ***p<0.001, ****p<0.0001.

In recipients of the XBB.1.5 monovalent vaccine, double-discrimination was used to define three populations of RBD-specific Bmem; WH1^+^ only (WH1^+^XBB.1.5^-^), WH1^+^XBB.1.5^+^ cross-reactive and XBB.1.5^+^ only (WH1^-^XBB.1.5^+^) (**Figure 2C**). In controls, the proportions of WH1^+^XBB.1.5^+^ and XBB.1.5^+^ only RBD-specific Bmem increased from pre to 1 month-post vaccination (**Figure 2D**). WH1 only Bmem did not expand, highlighting preferential boosting of XBB.1.5 RBD-specific Bmem as we have previously described.^30^ This pattern of preferential boosting did not occur in IBD patients, who did not display increased proportions of WH1^+^ only, WH1^+^XBB.1.5^+^ cross-reactive or XBB.1.5^+^ only Bmem, and had significantly lower proportions of these Bmem at 1 month post-vaccination than healthy controls (**Figure 2D**). Furthermore, absolute numbers of these three Bmem populations were similar in patients before and after vaccination (**Supplementary Figure 5B**). Between 1 and 6 months post-vaccination, controls had a significant decline in WH1^+^ only (0.05 vs 0.024 % overall Bmem, p=0.049), XBB.1.5^+^WH1^+^ (0.036 vs 0.011 % overall Bmem, p=0.0039) and XBB.1.5^+^ only RBD-specific Bmem (0.192 vs 0.072 % overall Bmem, p=0.002). This did not occur in IBD patients, who had an increase in XBB.1.5^+^ only RBD-specific Bmem from 1 to 6 month post-vaccination, suggesting that the preferential boosting of XBB.1.5 RBD-specific Bmem was delayed. GLMM analysis did not identify any demographic, vaccination or infection factors that impacted the differences observed between IBD patients and controls (data not shown). Concomitant treatment with immunomodulators had a significant effect on IBD patient RBD-specific Bmem frequency prior to vaccination (β: 0.04, P=0.05); however, no other patient-related factors were identified to significantly impact RBD-specific Bmem frequency post-vaccination.

### RBD-specific Bmem have altered activation and maturation in IBD patients

After finding lower frequencies of RBD-specific Bmem in IBD patients after vaccination, further phenotyping was performed to elucidate whether the activation profile and maturation status of these RBD-specific Bmem differed from controls. The frequencies of recently activated CD71^+^CD38^dim^ RBD-specific Bmem (**Figure 3A**) increased 1 month after vaccination in all recipients of either the WH1/BA.5 bivalent (IBD: 4.88 vs 10.23 % RBD-specific Bmem, p=0.001, control; 5.03 vs 10.7 % RBD-specific Bmem, p<0.0001) or XBB.1.5 monovalent vaccine (IBD: 4.35 vs 4.70 % RBD-specific Bmem, p=0.69, control; 7.56 vs 13.73 % RBD-specific Bmem, p=0.002) (**Figure 3B**). Following vaccination, CD71^+^CD38^dim^ RBD-specific Bmem frequencies were similar between IBD patients and controls, despite IBD patients having a significantly lower frequency than controls prior to vaccination. Similarly, the frequencies of CD21^lo^ RBD-specific Bmem (**Figure 3C**) increased in all vaccine recipients 1 month after vaccination, followed by a decline from 1 month to 6 months post-vaccination (**Figure 3D**). The frequencies of CD21^lo^ RBD-specific Bmem were significantly higher in IBD patients than controls both pre- and post-vaccination, a reflection of the phenotype observed in total Bmem (**Supplementary Figure 4B**).

**Figure 3.**
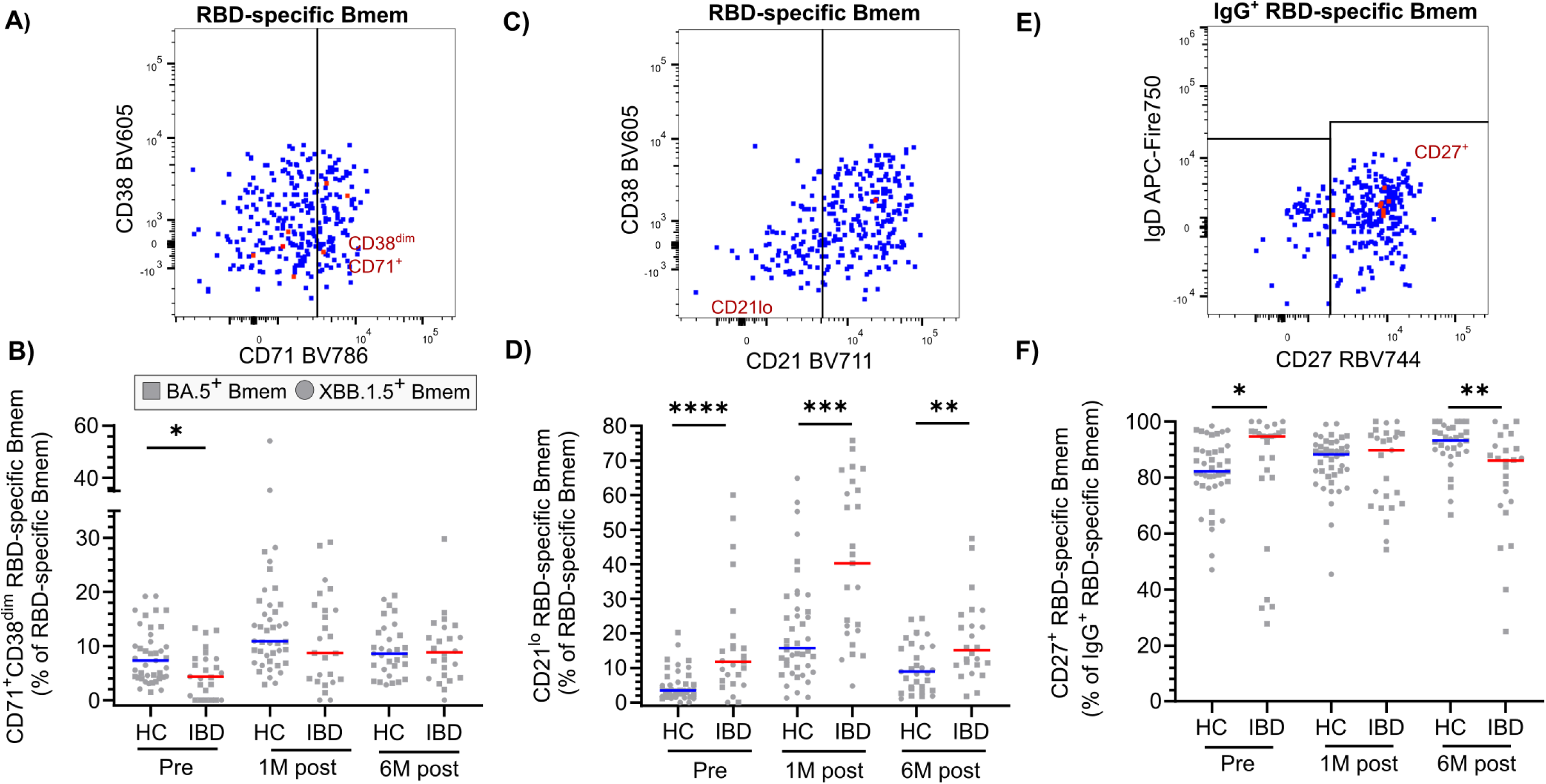
Phenotyping of RBD-specific Bmem in response to COVID-19 booster vaccination in IBD patients and controls. **(A)** Gating strategy to determine CD21^lo^ RBD-specific Bmem, **(B)** frequencies of CD21lo RBD-specific Bmem as a proportion of RBD-specific Bmem, **(C)** gating strategy to determine CD71^+^CD38^dim^ RBD-specific Bmem, **(D)** frequencies of CD71^+^CD38^dim^ RBD-specific Bmem as a proportion of RBD-specific Bmem, **(E)** Gating strategy to determine CD27+ cells withWH1/in IgG+ RBD-specific Bmem, **(F)** frequencies of CD27^+^ within IgG+ RBD-specific Bmem. Horizontal lines depict medians. Squares represent BA.5 RBD-specific Bmem from BA.5 bivalent vaccinated individuals (n=19 IBD patients, n=22 controls), Circles represent XBB.1.5 RBD-specific Bmem from XBB.1.5 monovalent vaccinated individuals (n=7 IBD patients, n=22 controls). Statistical significance between patients and controls as calculated by Mann-Whitney test. *p<0.05, **p<0.01, ***p<0.001, ****p<0.0001.

The proportion of CD27^+^ cells within IgG^+^ RBD-specific Bmem were assessed as a marker for secondary GC-experienced class-switched Bmem (**Figure 3E**).^46^ The frequencies of CD27^+^IgG^+^ RBD-specific Bmem in controls significantly increased (82.1 vs 88.3 % RBD-specific Bmem, p=0.002) after vaccination (**Figure 3F**). In contrast, the frequencies of CD27^+^IgG^+^ RBD-specific Bmem in IBD patients decreased after vaccination, and at 6 months post-vaccination, these were significantly lower than in controls. This pattern appeared to be independent of the total Bmem population, in which the frequencies of CD27^+^IgG^+^ Bmem were similar pre- and post-vaccination (**Supplementary Figure 4C**).

### Altered Ig isotype and IgG subclass distributions in RBD-specific Bmem

Next, the Ig isotype and IgG subclass distributions of BA.5 and XBB.1.5 RBD-binding Bmem were determined pre-vaccination, and 1 and 6 months post-booster vaccination (**Figure 4A**). In all vaccine recipients, the vast majority of BA.5 and XBB.1.5 RBD-specific Bmem expressed IgG1, which expanded 1 month post-vaccination and then declined from 1 month to 6 months post-vaccination (**Figure 4B,C**). As IBD patients had significantly fewer RBD-specific Bmem following vaccination than controls **(Figure 2B,D)** the proportions of IgG1^+^ RBD-specific Bmem within total Bmem were lower pre and post-vaccination (**Figure 4C**). The change in Ig isotype distribution from 1 to 6 months post-vaccination was also driven by increased proportions of IgD^+^IgM^+^ RBD-specific Bmem within total Bmem at 6 months (**Figure 4B,D**). IgD^+^IgM^+^ RBD-specific Bmem were highest in IBD patients who received the XBB.1.5 monovalent vaccine **(Figure 4D)**.

**Figure 4.**
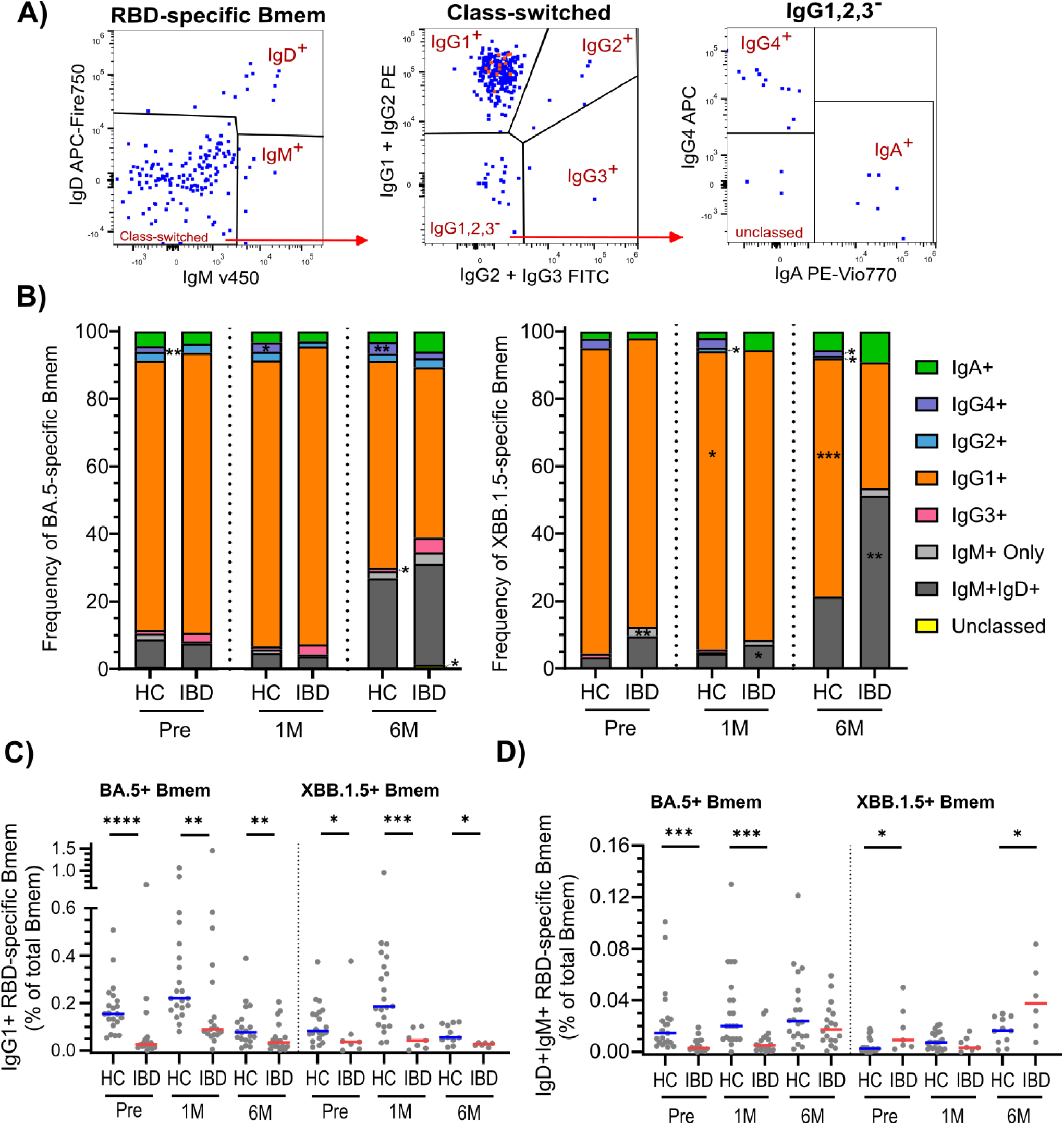
Immunoglobulin isotype and IgG subclass expression by RBD-specific Bmem. **(A)** Gating strategy to determine Ig Isotype and IgG subclass expression on RBD-specific Bmem **(B)** Distribution of Immunoglobulin isotype and IgG subclass expressing subsets within RBD-specific Bmem from WH1/BA.5 bivalent (n=19 patients, n=22 controls) and XBB.1.5 monovalent vaccine recipients (n=7 IBD patients, n=22 controls). **(C)** Frequency of IgG1^+^ RBD-specific Bmem as proportion of total Bmem, **(D)** Frequency of IgD^+^IgM^+^ RBD-specific Bmem as proportion of total Bmem. Statistical significance of frequencies between patients and controls as calculated by Mann-Whitney test. *p<0.05, **p<0.01, ***p<0.001.

Differences in the expression of IgG subclasses within RBD-specific Bmem between IBD patients and controls were similar to that observed in total Bmem. Indeed, the frequencies of IgG4^+^BA.5^+^ RBD-specific Bmem were significantly lower in IBD patients than in controls who received WH1/BA.5 bivalent vaccination (**Figure 4B**). Furthermore, the frequencies of IgG3^+^BA.5^+^ RBD-specific Bmem were higher in IBD patients than controls (**Figure 4B**). A similar trend was also seen in patients who received the XBB.1.5 monovalent vaccine, with the frequencies of IgG4^+^XBB.1.5^+^ RBD-specific Bmem significantly lower in IBD patients than controls post-vaccination. In addition, IgG2^+^XBB.1.5^+^ RBD-specific Bmem were also significantly lower in patients than controls 6 months after receiving the XBB.1.5 monovalent vaccine (**Figure 4B**).

### Reduced variant-binding capacity of antibodies in IBD patients after booster vaccination

As the SARS-CoV-2 spike RBD continued to evolve antigenically, we next examined the cross-reactive potential of antibodies generated in response to vaccination. IBD patients that received the WH1/BA.5 bivalent vaccine had significantly higher levels of BA.1 (12.1 vs 27.9 µg/mL, p=0.0004), XBB.1.5 (12.0 vs 29.6 µg/mL, p<0.0001), and JN.1 (2.28 vs 10.3 µg/mL, p p<0.0001) RBD-binding IgG after vaccination (**Figure 5A**). However, BA.1 and JN.1 RBD-binding IgG were significantly lower at 1 month post-vaccination in IBD patients than in controls. Moreover, IBD patients also had significantly lower levels of BA.1, XBB.1.5, and JN.1 RBD-binding IgG than controls at 6 months post-vaccination.

**Figure 5.**
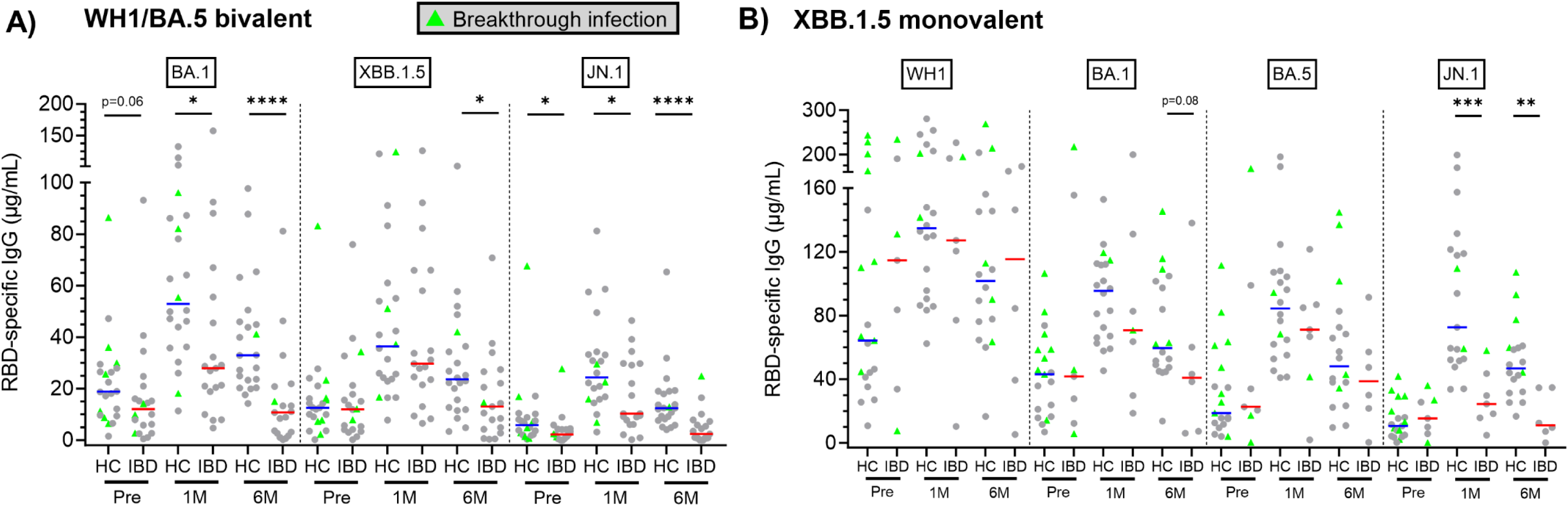
Variant RBD-specific IgG levels before and after COVID-19 booster vaccination in IBD patients and controls. **(A)** concentrations of BA.1, XBB.1.5 and JN.1 RBD-specific IgG from individuals who received BA.5 bivalent vaccination (n=19 IBD patients, n=22 controls), **(B)** concentrations of WH1, BA.1, BA.5 and JN.1 RBD-specific IgG from individuals who received XBB.1.5 monovalent vaccination (n=7 IBD patients, n=22 controls). Data shown as median. Statistical significance between IBD patients and controls as calculated by Mann-Whitney test. Breakthrough infection as determined by seropositivity to SARS-CoV-2 Nucleoprotein (NCP). ****p<0.0001, ***p<0.001, **p<0.01, *p<0.05.

Following XBB.1.5 monovalent vaccination, IBD patients and controls had comparable levels of WH1, BA.1 and BA.5 RBD-specific IgG **(Figure 5B)**. However, IBD patients had significantly lower JN.1 RBD-binding IgG after vaccination, highlighting potentially impaired ability to respond to emerging variants. GLMM analysis highlighted that variant-binding antibody levels at 1 month (β: 0.22, p=0.010) and 6 months (β: 0.20, p=0.018) post-vaccination were positively associated with the number of previously-received vaccine doses. Furthermore, breakthrough infection in the period between 1 month to 6 months post-vaccination was positively associated with total variant-binding antibody levels at 6 months (β: 0.36, p=0.047). We observed a similar pattern of reduced NAb titers against additional variants 6 months post-vaccination in IBD patients who received the XBB.1.5 monovalent vaccine (**Supplementary Figure 6**). Taken together, this highlights that the durability of variant-binding antibodies is poorer in IBD patients, which may be improved by additional booster vaccinations.

### Lower variant cross-recognition by RBD-specific Bmem from IBD patients

Having observed that antibody responses in IBD patients have a reduced capacity to cross-react to other SARS-CoV-2 variants, we assessed additional non-vaccine variant binding by their RBD-specific Bmem. Due to the more recent emergence of the JN.1 variant, this variant was only examined in 2 controls pre and 1 month post-WH1/BA.5 vaccination. BA.5 RBD-specific Bmem from all recipients that received the WH1/BA.5 bivalent vaccine had a significant increase in binding to WH1 (IBD: 0.02 vs 0.07 % overall Bmem, p=0.0008, control; 0.07 vs 0.14 % overall Bmem, p<0.0001), BA.1 (IBD: 0.02 vs 0.08 % overall Bmem, p=0.001, control; 0.11 vs 0.16 % overall Bmem, p<0.0001), XBB.1.5 (IBD: 0.03 vs 0.09 % overall Bmem, p=0.001, control; 0.03 vs 0.05 % overall Bmem, p=0.005) and JN.1 (IBD: 0.02 vs 0.06 % overall Bmem, p=0.002) RBD at 1 month post-vaccination (**Figure 6A,B, Supplementary Figure 7A,B**). However, the frequencies of BA.5 RBD-specific Bmem that could bind non-vaccine variants were significantly lower in patients than in controls. Examining the kinetics of variant-binding by BA.5 RBD-specific Bmem before and after WH1/BA.5 bivalent vaccination revealed that controls had a more pronounced increase in binding to WH1 and BA.1 variants than IBD patients at 1 month post-vaccination (**Figure 6C**). The frequencies of BA.5 RBD-specific Bmem co-binding additional variants declined from 1 month to 6 months post-WH1/BA.5 bivalent vaccination, with IBD patients exhibiting significantly lower binding to all variants than controls 6 months post-vaccination (**Figure 6B,C**).

**Figure 6.**
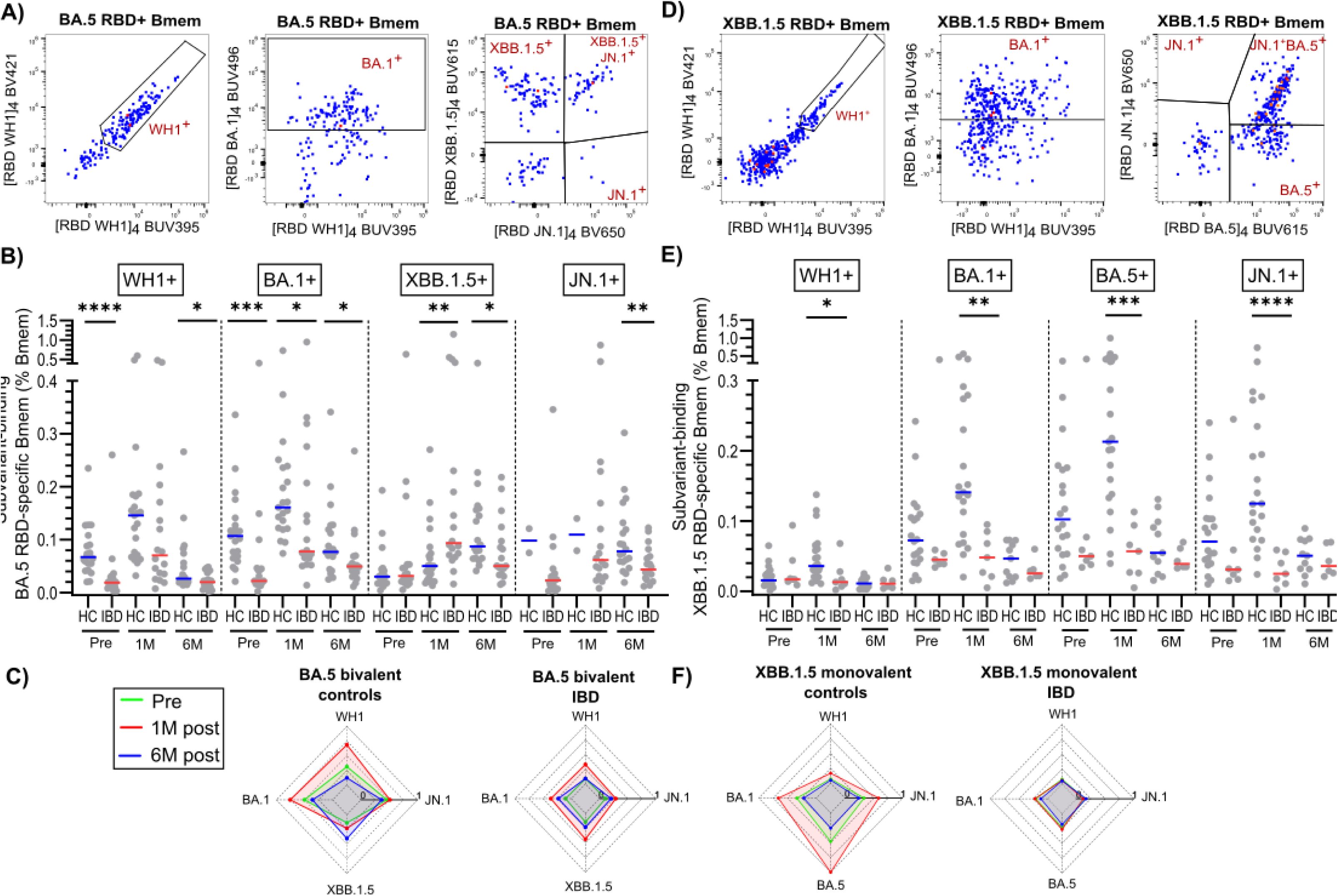
Variant cross-binding of RBD-specific Bmem before and after COVID-19 booster vaccination in IBD patients and controls. **(A)** Gating strategy to determine cross-binding to WH1, BA.1, XBB.1.5 and JN.1 within BA.5+ RBD-specific Bmem. **(B)** Frequencies of BA.5 RBD-specific Bmem cross-binding to WH1, BA.1, XBB.1.5 and JN.1 in WH1/BA.5 bivalent vaccine recipients (n=19 IBD patients, n=22 controls), **(C)** radar plots of median relative RBD-specific variant binding from controls and IBD patients before and after BA.5 bivalent vaccination. **(D)** Gating strategy to determine cross-binding to WH1, BA.1, BA.5 and JN.1 within XBB.1.5+ RBD-specific Bmem. **(E)** Frequencies of XBB.1.5 RBD-specific Bmem cross-binding to WH1, BA.1, BA.5 and JN.1 in XBB.1.5 monovalent vaccinated individuals (n=7 IBD patients, n=22 controls), **(F)** radar plots of median relative RBD-specific variant binding from controls and IBD patients before and after XBB.1.5 monovalent vaccination. Statistical significance between patients and controls as calculated by Mann-Whitney test. *p<0.05, **p<0.01, ***p<0.001.

After XBB.1.5 monovalent vaccination, the frequencies of XBB.1.5 RBD-specific Bmem that could also bind WH1, BA.1, BA.5, and JN.1 increased in controls **(Figure 6D,E)**; however, this did not occur in IBD patients. Consequently, XBB.1.5 RBD-specific Bmem from IBD patients had significantly lower frequencies of variant binding at 1 month post-vaccination than healthy controls (**Figure 6E**). This reduced binding was more pronounced than observed following WH1/BA.5 bivalent vaccination (**Figure 6F**). GLMM analysis highlighted that IBD patients receiving infliximab monotherapy had significantly higher total variant cross-binding by RBD-specific Bmem than those on combination therapy pre-vaccination (β: 0.04 p=0.048); however this did not impact responses post-vaccination.

## DISCUSSION

The immunocompetency of IBD patients on anti-TNF biologics following immunization with COVID-19 booster formulations is poorly defined, potentially contributing to a lack of evidence-based vaccination recommendations for patients. In the current study, we show that IBD patients on anti-TNF biologics mount antibody responses and form vaccine-specific Bmem following WH1/BA.5 bivalent or XBB.1.5 monovalent mRNA COVID-19 booster vaccination. However, patients’ antibody responses and Bmem numbers were lower than in controls at 1- and 6 months after vaccination. Furthermore, RBD-specific antibodies and Bmem from patients had a reduced capacity to recognize multiple SARS-CoV-2 Omicron subvariants. Taken together, this highlights that IBD patients treated with anti-TNF biologics display impaired immune memory formation.

mRNA-based COVID-19 booster vaccinations continue to provide protection against severe COVID-19 in the general population.^47–49^ We found that IBD patients on anti-TNF therapy form antibody and Bmem responses to both WH1/BA.5 bivalent and XBB.1.5 monovalent vaccines, with detectable antibodies and RBD-specific Bmem 6 months post-vaccination, indicative of ongoing humoral immune protection. Still, both RBD-specific IgG and NAb titers decreased strongly from 1 month to 6 months post-vaccination across both IBD patients and controls, and RBD-specific IgG from IBD patients on anti-TNF exhibited reduced antibody durability compared to controls, as previously observed for the primary double-dose vaccination.^13,50^ These observations fit with estimated vaccine efficacy against symptomatic infection for the XBB.1.5 monovalent vaccine, which is in the range of 47.6-57.6% for healthy individuals, with rapid waning of protection within months of vaccination.^47^ In contrast, XBB.1.5 booster vaccination efficacy against hospitalization in a broad range of immunocompromised patients has been estimated at 23%, likely as a consequence of the poorer durability of RBD-specific antibodies.^51^

The rapid decline in serum antibodies in IBD patients is likely a consequence of anti-TNF treatment reducing generation of long-lived GC-derived plasma cells that produce long-lasting IgG.^52^ Interestingly, we observed that IBD patients’ antibody responses 1 month after booster vaccination were more comparable to controls, suggesting that the response from short-lived plasma cells derived from extrafollicular sites was similar between patients and controls. Further profiling of glycosylation patterns on RBD-specific IgG may help to determine the source of these antibodies. Long-lasting GC-derived antibodies display low galactosylation and sialylation, although these patterns may also be impacted by inflammation in IBD.^53,54^

We observed differences in the kinetics of responses between vaccine antigens, with higher durability of antibody and Bmem after XBB.1.5 monovalent vaccination than after BA.5 bivalent vaccination in patients. Although monovalent and bivalent vaccines contain the same total amount of antigen-coding mRNA, this is split evenly between WH1 and BA.5 antigens in the bivalent vaccine. Consequently, the XBB.1.5 monovalent formulation contains twice the amount of mRNA encoding the XBB.1.5 antigen.^55,56^ This may partially explain the improved durability seen in IBD patients, as primary vaccination with the higher-dose Moderna mRNA-1273 was associated with increased RBD-specific antibody titers 6 months after vaccination in healthy individuals when compared to the lower-dose Pfizer BNT162b2 vaccine.^57,58^ Further, GLMM analyses revealed that the number of vaccine doses previously received was a significant predictor for antibody levels 6 months post-booster vaccination. Together, these observations support the need for continued booster vaccine uptake in IBD patients with anti-TNF treatment, to maintain adequate protection against SARS-CoV-2 infection.

Following a booster with variant-containing vaccines, Bmem from IBD patients on anti-TNF showed enhanced Omicron subvariant recognition, overcoming potential effects of immune imprinting. Immune imprinting caused by pre-existing immune memory specific for the SARS-CoV-2 ancestral (WH1) strain was thought to limit naïve B cells from being exposed to subsequent variant antigens, potentially reducing the Bmem response to emerging SARS-CoV-2 variants.^59,60^ Despite reduced RBD-specific Bmem in IBD patients prior to vaccination, booster vaccination could still boost cross-reactive Bmem that recognize antigens contained in the vaccine, likely derived from the pre-existing pool of RBD-specific Bmem.^61^ However, the cross-reactivity of the response appears limited compared to controls. Given that no differences in the naïve B cell pool were seen between our patients and controls, this is in line with previous observations that the breadth of variant-binding is dependent on the diversity of the pre-existing Bmem pool.^62^ Indeed, natural infection is also likely to influence the B-cell receptor repertoire within the pool of RBD-specific Bmem, although GLMM did not identify previous infection to be a significant factor impacting cross-reactivity of Bmem.^63^ Despite poorer non-vaccine variant-binding capacity of RBD-specific antibodies and Bmem in patients, we still observed increased antibodies against JN.1 after vaccination, similar to another study of IBD patients.^31^ This supports the recommendation for regular repeated COVID-19 booster doses with updated variant formulations in order to maintain a pool of cross-reactive Bmem that can respond to upcoming emerging variants.

IBD patient RBD-specific Bmem displayed different phenotypic kinetics in response to COVID-19 booster vaccination compared to controls. CD27 expression was altered after vaccination, with significantly fewer CD27^+^ IgG^+^ RBD-specific Bmem in patients 6 months after vaccination compared to controls. This is likely a consequence of impaired GC responses caused by ongoing anti-TNF treatment. TNF-α plays a critical role in the formation and functions of the GC response, particularly in the maintenance of follicular dendritic cells and GC B cells.^14^ Moreover, IBD patients also have reduced circulating follicular helper T cells shortly after COVID-19 vaccination, strongly correlating with reduced RBD-specific antibodies and Bmem.^50^

The impaired GC response is likely to have contributed to altered Ig isotype and IgG subclass distributions we observed in Bmem in IBD patients. IBD patients had fewer IgG4^+^ and IgG1^+^ RBD-specific Bmem, likely reflective of impaired class switching that has previously been observed to occur after initial COVID-19 vaccination in IBD patients on anti-TNF.^32^ Persisting IgG4^+^ RBD-specific Bmem observed after primary course of ancestral mRNA-based COVID-19 vaccination in healthy individuals has been proposed to be a consequence of an ongoing GC reaction resulting in consecutive class-switch recombination from proximal IgG3 to distal IgG4 along the four gamma heavy-chain genes within the Ig gene complex^19,64^. Interestingly, we observed that the frequency of IgG3^+^ RBD-specific Bmem was also increased in patients which may be reflective of reduced GC responses over time. In contrast, we did not observe any differences in the pattern of CD71 expression in patients after vaccination, highlighting that some aspects of the Bmem response to vaccination are not impaired by anti-TNF treatment.^19,65^ Indeed, the T-cell response in patients receiving anti-TNF is not impaired. Spike-specific CD4^+^ and CD8^+^ T cells maintain equivalent durability to healthy individuals up to 6 months after both primary vaccination and after booster.^12,66–68^ The T-cell response may even be enhanced in immunocompromised people as a compensatory mechanism.^69–71^

Furthermore, we observed that IBD patients had a higher frequency of CD21^lo^ Bmem, likely reflective of the inflammatory environment previously observed in patients with IBD and other autoimmune diseases.^45,72^ These cells have previously been shown to express CD11c, and together with a downregulation of CD27 may represent a pool of atypical memory B cells that may be predominantly derived from extrafollicular responses.^73^ These responses appear to be larger in IBD patients, as we also observed that the total plasmablast pool was consistently higher in IBD patients across the entire study when compared to controls. SARS-CoV-2 spike-specific plasmablasts have previously been observed to be increased in IBD patients on anti-TNF after primary COVID-19 vaccination.^50^ This suggests an enhanced extrafollicular responses may be an ongoing consequence of anti-TNF treatment, impacting RBD-specific Bmem formation even after several COVID-19 booster vaccinations. This may also impact responses to other vaccines. Influenza specific IgG levels are also lower in IBD patients receiving anti-TNF when compared to healthy controls.^74^

Although our IBD patient cohort is larger than previous studies that have examined RBD-specific Bmem,^12,32,50^ it is important to acknowledge that our study may be underpowered to examine additional clinical factors that impact vaccine immunogenicity. Previous studies of COVID-19 primary vaccination have highlighted that concomitant immunomodulator or steroid use are associated with reduced RBD-specific antibodies in anti-TNF treated IBD patients.^10, 17,75^ Although 56% of patients in our study cohort were on concomitant immunomodulating drugs, none were receiving steroids. We did not find any significant difference in the serological response to vaccination between combination or monotherapy, although combination therapy significantly impacted RBD-specific Bmem frequencies pre-vaccination. Furthermore, we did not recruit IBD patients on other biologics, such as vedolizumab or ustekinumab, as these have been previously shown to have less impact on the serological response to vaccination.^75^ These patients may have been useful to delineate the impact of anti-TNF treatment from disease on the phenotype of Bmem, particularly as infliximab may partially restore abnormalities in Ig isotype switching seen in Crohn’s disease patients.^45^ Healthy control samples used in our study were matched based on the booster vaccines received by patients. Although demographic, vaccination and infection history were factored into our GLMM analysis, we found that natural infection did significantly impact RBD-specific antibody levels, particularly at baseline.

Within the current context of variable vaccination recommendations around the world, our data represents the most detailed profiling of immunocompetency in IBD patients receiving anti-TNF treatment. We report that patients mount a serological and Bmem response to vaccination; however, the capacity, durability and cross-reactivity of these responses are lower than controls. Consequently, these findings of impaired immune memory suggest that IBD patients receiving anti-TNF remain at risk of infection, even after vaccination. Hence, patients with this iatrogenic immunodeficiency will benefit from ongoing annual booster vaccination to prevent symptomatic breakthrough infections and pathogen spread.

## Acknowledgements

We would like to thank Dr. Jack Edwards, Ms. Shir Sun, Ms. Pei Aui and the Alfred Hospital Medical day unit staff for their assistance with collection and processing of study blood samples. We also thank the ARA flow core facility staff for support with Flow Cytometry.

## Author Contributions

Conceptualization: MCvZ, MS, REOH, HED; Patient recruitment: JH, KD, JM, RDdV; Data collection and experimental work: PG, LB, AW, IB, HAF, DG, LMZ; Data analyses: LB, AW, IB, PG, HED; Manuscript drafting: PG, MCvZ; Overall supervision: MCvZ, PG, MS, PMH, RDdV, HED. All authors read, edited and approved the final manuscript.

## Data availability

The data underlying this article will be shared on reasonable request to the corresponding author.

## Abbreviations

Bmem: memory B cell
NAb: neutralizing antibody
IgG: immunoglobulin G
IBD: inflammatory bowel disease
UC: ulcerative colitis
CD: Crohn’s disease
TNF-α: tumor necrosis factor α
RBD: receptor binding domain
BCR: B cell receptor
GC: germinal center

