## Supplementary materials for "Impaired memory B-cell formation after mRNA-based COVID-19 booster vaccination in patients with inflammatory bowel disease receiving anti-TNF treatment"

### Supplemental Tables and Figures

#### Supplementary Tables (n=3)

#### Supplementary Table 1. Antibody panel compositions

| | **Fluorochrome** | | | | | | | | | | | | | | | | | | | |
|---|---|---|---|---|---|---|---|---|---|---|---|---|---|---|---|---|---|---|---|---|
| **Tube** | BUV 395 | UV 450 | BUV 496 | BUV 615 | BUV 737 | BUV 805 | BV 421 | V450 | BV480 | BV605 | BV650 | BV711 | BV786 | FITC | RB744/ PerCP Cy-5.5 | PE | PE-Vio670 | BYG 710 | PE-Cy7/ PE-Vio770 | APC | APC-Cy7/ APC-Fire750 |
| **1. Trucount[a]** | — | — | — | — | — | — | — | — | — | — | — | — | — | CD3 | CD45 | CD16+ CD56 | — | — | CD4 | CD19 | CD8a |
| **2a. Bmem (BA.5 bivalent recipients)** | WH1 RBD | Fixable viability | BA.1 RBD | XBB.1.5 RBD | BA.5 RBD | CD3 | WH1 RBD | IgM | BA.5 RBD | CD38 | JN.1 RBD[b] | CD21 | CD71 | IgG2+ IgG3 | CD27 | IgG1+ IgG2 | CD11c[c] | CD19 | IgA | IgG4 | IgD |
| **2b. Bmem (XBB.1.5 monovalent recipients)** | WH1 RBD | Fixable viability | BA.1 RBD | BA.5 RBD | XBB.1.5 RBD | CD3 | WH1 RBD | IgM | XBB.1.5 RBD | CD38 | JN.1 RBD | CD21 | CD71 | IgG2+ IgG3 | CD27 | IgG1+ IgG2 | CD11c[c] | CD19 | IgA | IgG4 | IgD |
| **3. Strep control** | Strep | Fixable viability | Strep | — | Strep | CD3 | Strep | — | Strep | — | Strep | — | — | — | CD27 | — | — | CD19 | — | — | IgD |

[a]Only run for 23 of 41 BA.5 bivalent recipients (n=19 IBD patients, n=4 healthy controls) and 9 of 29 XBB.1.5 recipients (n=6 IBD patients, n=3 heathy controls) across all 3 timepoints.

[b]Only run for 21 of 41 BA.5 bivalent recipients (n=19 IBD patients, n=2 healthy controls) and 14 of 29 XBB.1.5 recipients (n=6 IBD patients, n=8 heathy controls) across all 3 timepoints.

[c]Only run for 15 of 41 BA.5 bivalent recipients (n=12 IBD patients, n=3 healthy controls) and 9 of 29 XBB.1.5 recipients (n=6 IBD patients, n= 3 healthy controls) across all 3 timepoints

**Supplementary Table 2. Antibody details**

| Marker | Fluorochrome | Clone | Vendor | Cat. number | Volume (µL)/ 100µL test | Tube |
|---|---|---|---|---|---|---|
| CD3 | BUV805 | UCHT1 | BD Biosciences | 612896 | 2.5 | 2,3 |
| CD3 | FITC | SK7 | BD Biosciences | 662995* | 46 ng | 1 |
| CD4 | PE-Cy7 | SK3 | BD Biosciences | 662995* | 30 ng | 1 |
| CD8a | APC-Cy7 | SK1 | BD Biosciences | 662995* | 126 ng | 1 |
| CD16 | PE | B73.1 | BD Biosciences | 662995* | 33 ng | 1 |
| CD19 | APC | SJ25C1 | BD Biosciences | 662995* | 46 ng | 1 |
| CD19 | BYG710 | HIB19 | Cytek Biosciences | SKU-R7-20010 | 1 | 2,3 |
| CD11c | PE-Vio770 | REA618 | Miltenyi Biotec | 130-133-819 | 0.4 | 2 |
| CD21 | BV711 | B-ly4 | BD Biosciences | 563163 | 2.5 | 2 |
| CD27 | RB744 | M-T271 | BD Biosciences | 570713 | 2.5 | 2,3 |
| CD38 | BV605 | HB7 | BD Biosciences | 562665 | 0.2 | 2 |
| CD45 | PerCP Cy-5.5 | 2D1 | BD Biosciences | 662995* | 120 ng | 1 |
| CD56 | PE | NCAM16.2 | BD Biosciences | 662995* | 22 ng | 1 |
| CD71 | BV786 | M-A712 | BD Biosciences | 563768 | 1 | 2 |
| Fixable Viability | Live dead blue | - | Invitrogen | L34961 | 0.1 | 2,3 |
| IgA | PE-Vio770 | REA1014 | Miltenyi Biotec | 130-117-003 | 1 | 2 |
| IgD | APC-Fire750 | W18340F | Biolegend | 307822 | 0.2 | 2,3 |
| IgG1 | PE | G17-1 | BD Biosciences | 624049 | 0.1 | 2 |
| IgG2 | FITC | HP6002 | BD Biosciences | 624045 | 0.5 | 2 |
| IgG2 | PE | HP6002 | BD Biosciences | 624049 | 1 | 2 |
| IgG3 | FITC | HP6047 | BD Biosciences | 624045 | 0.5 | 2 |
| IgG4 | APC | SAG4 | Cytognos | CYT-IGG4AP | 2 | 2 |
| IgM | Hor V450 | MHM88 | Cytek Biosciences | Custom | 0.25 | 2 |
| Streptavidin | BUV395 | - | BD Biosciences | 564176 | 0.67 | 2,3 |
| Streptavidin | BUV496 | - | BD Biosciences | 612961 | 0.67 | 2,3 |
| Streptavidin | BUV615 | - | BD Biosciences | 613013 | 0.67 | 2,3 |
| Streptavidin | BUV737 | - | BD Biosciences | 612775 | 0.67 | 2,3 |
| Streptavidin | BV421 | - | BioLegend | 405225 | 0.13 | 2,3 |
| Streptavidin | BV480 | - | BD Biosciences | 564876 | 0.67 | 2,3 |
| Streptavidin | BV650 | - | BioLegend | 563855 | 0.13 | 2,3 |

*Antibodies part of the Multitest™ 6-color TBNK kit (BD Biosciences, Cat. number 662967)

**Supplementary Table 3. Patient characteristics**

| Patient Number | Age range at Recruitment (years) | Sex (M/F) | Clinical Diagnosis | Vaccine (variant) | Blood Sampling Timing (Days) | | NCP Seropositive during study (-/+) | IV Infliximab Treatment Course | Serum Infliximab level (µg/mL) | Concomitant Immunomodulators |
|---|---|---|---|---|---|---|---|---|---|---|
| | | | | | 1 M Post | 6 M Post | | | | |
| **1** | 41-50 | F | UC | BNT162b2 (BA.4/5) | 30 | 184 | - | 10 mg/kg, 4 weekly | 13.8 | Azathioprine (100 mg/d) |
| **2** | 21-30 | M | CrD | BNT162b2 (BA.4/5) | 21 | 231 | - | 5 mg/kg, 6 weekly | 9.65 | Azathioprine (200 mg/d) |
| **3** | 21-30 | M | CrD | mRNA-1273.222 (BA.4/5) | 51 | 92 | - | 5 mg/kg, 6 weekly | 24.8 | Azathioprine (200 mg/d) |
| **4** | 51-60 | F | CrD | BNT162b2 (BA.4/5) | 28 | 196 | - | 5 mg/kg, 8 weekly | 9.29 | - |
| **5** | 41-50 | M | CrD | BNT162b2 (BA.4/5) | 30 | 210 | + | 5 mg/kg, 8 weekly | 14.5 | - |
| **6** | 21-30 | M | CrD | mRNA-1273.214 (BA.1) | 23 | 224 | + | 5 mg/kg, 6 weekly | 9.49 | Methotrexate (10 mg weekly) |
| **7** | 21-30 | F | CrD | BNT162b2 (BA.4/5) | 20 | 223 | - | 10 mg/kg, 4 weekly | 2.91 | Azathioprine (25 mg/d) |
| **8** | 41-50 | M | UC | mRNA-1273.222 (BA.4/5) | 26 | 173 | - | 10 mg/kg, 4 weekly | 23.9 | - |
| **9** | 31-40 | F | CrD | BNT162b2 (BA.4/5) | 16 | 184 | - | 5 mg/kg, 8 weekly | 3.59 | - |
| **10** | 21-30 | M | CrD | mRNA-1273.222 (BA.4/5) | 29’ | 218 | - | 10 mg/kg 6 weekly | 16.3 | Azathioprine 50 mg |
| **11** | 51-60 | M | CrD | mRNA-1273.222 (BA.4/5) | 24 | 188 | - | Infliximab (5 mg/kg, 6 weekly) | 16.6 | - |
| **12** | 71-80 | F | UC | mRNA-1273.222 (BA.4/5) | 51 | 219 | - | 10 mg/kg, 8 weekly | 24.4 | - |
| **13** | 51-60 | M | UC | BNT162b2 (BA.4/5) | 23 | 219 | - | 5 mg/kg, 8 weekly | No data | - |

| Patient Number | Age range at Recruitment (years) | Sex (M/F) | Clinical Diagnosis | Vaccine (variant) | Blood Sampling Timing (Days) | | NCP Seropositive during study (-/+) | IV Infliximab Treatment Course | Serum Infliximab level (µg/mL) | Concomitant Immunomodulators |
|---|---|---|---|---|---|---|---|---|---|---|
| | | | | | 1 M Post | 6 M Post | | | | |
| **14** | 21-30 | M | CrD | BNT162b2 (BA.4/5) | 28 | 210 | - | 8 mg/kg, 8 weekly | 12.5 | Methotrexate (10 mg weekly) |
| **15** | 21-30 | F | UC | BNT162b2 (BA.4/5) | 23 | 237 | + | 5 mg/kg, 8 weekly | 35.7 | Azathioprine (150 mg/d) |
| **16** | 31-40 | M | UC | mRNA-1273.222 (BA.4/5) | 40 | 244 | + | 5mg/kg, 6 weekly | 3.17 | Mercaptopurine (75 mg) |
| **17** | 21-30 | F | UC | BNT162b2 (BA.4/5) | 31 | 214 | - | 5 mg/kg, 8 weekly | 6.81 | - |
| **18** | 41-50 | M | CrD | BNT162b2 (BA.4/5) | 28 | 204 | - | 5 mg/kg, 6 weekly | 9.54 | Thioguanine (20 mg/d) |
| **19** | 31-40 | M | UC | mRNA-1273.222 (BA.4/5) | 57 | 169 | - | 10 mg/kg, 4 weekly | 35.5 | Methotrexate (10 mg/weekly) |
| **20** | 21-30 | M | CrD | BNT162b2 (XBB.1.5) | 61 | 203 | + | 5mg/kg, 8 weekly | No data | Methotrexate (12.5 mg/week) |
| **21** | 31-40 | F | CrD | mRNA-1273.222 (BA.4/5) | 30 | 198 | - | 5 mg/kg, 8 weekly | tbc | - |
| **22** | 41-50 | M | CrD | BNT162b2 (XBB.1.5) | 66 | n/a | + | 10 mg/kg, 8 weekly | 21.6 | - |
| **23** | 31-40 | F | CrD | BNT162b2 (XBB.1.5) | 31 | 170 | + | 5 mg/kg, 6 weekly | 20.2 | Mercaptopurine 25 mg/d Allopurinol (10 mg/d) |
| **24** | 31-40 | M | CrD | mRNA-1273.815 (XBB.1.5) | 32 | 130 | - | 5 mg/kg, 8 weekly | 1.84 | - |
| **25** | 41-50 | F | CrD | mRNA-1273.815 (XBB.1.5) | 70 | 154 | - | 10 mg/kg, 6 weekly | no data | Azathioprine |
| **26** | 71-80 | F | CrD | BNT162b2 (XBB.1.5) | 67 | 153 | - | 10 mg/kg, 4 weekly | no data | Mercaptopurine |
| **27** | 51-60 | M | UC | BNT162b2 (XBB.1.5) | 44 | 182 | + | 5 mg/kg, 8 weekly | no data | - |

IV: Intravenous, CrD: Crohn’s Disease, UC: Ulcerative colitis, NCP: Nucleocapsid protein

Supplementary Figures (n=7)

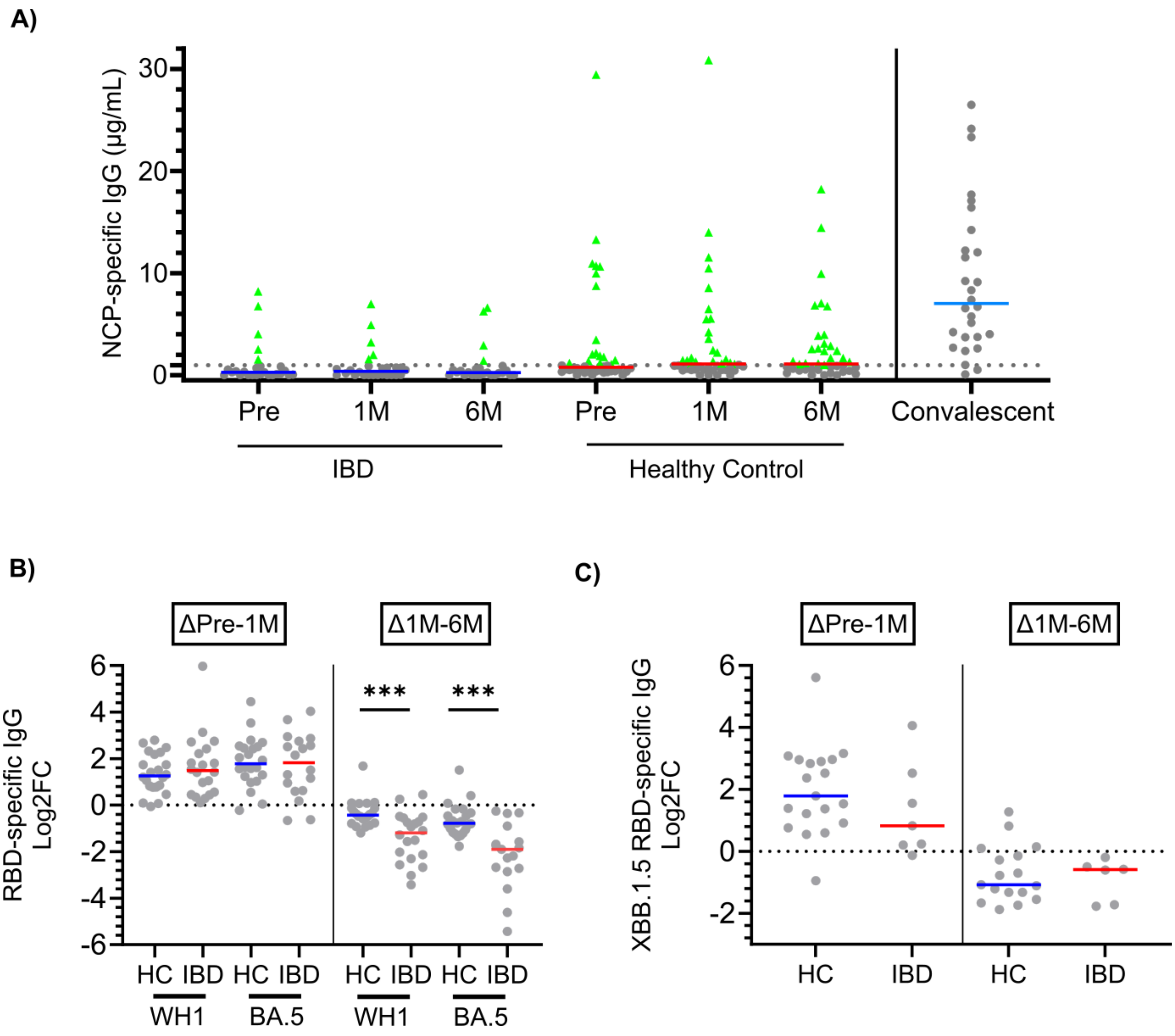


**Supplementary Figure 1. Nucleocapsid serology and kinetics of RBD-specific serum IgG before and after COVID-19 booster vaccination. (A)** Levels of Nucleocapsid (NCP)-specific IgG in IBD patients (n=27) and controls (HC, n=44) before and after COVID-19 booster vaccination. NCP seropositivity cut-off was determined from convalescent cohort published in Hartley *et al.*(39) **(B)** Log2 Fold-changes (FC) in WH1 and BA.5 RBD-specific IgG concentrations between: baseline and 1 month post, and 1 and 6 months post-vaccination in IBD patients (n=19) and controls (n=22) who received WH1/BA.5 bivalent vaccination. **(C)** Log2 Fold-changes (FC) in XBB.1.5 RBD-specific IgG concentrations between: baseline and 1 month post-vaccination, and 1 and 6 months post-vaccination in IBD patients (n=7) and controls (n=22) who received XBB.1.5 monovalent vaccination. Data shown as median. Statistical significance between patients and controls as calculated by Mann-Whitney test. ***p<0.001.

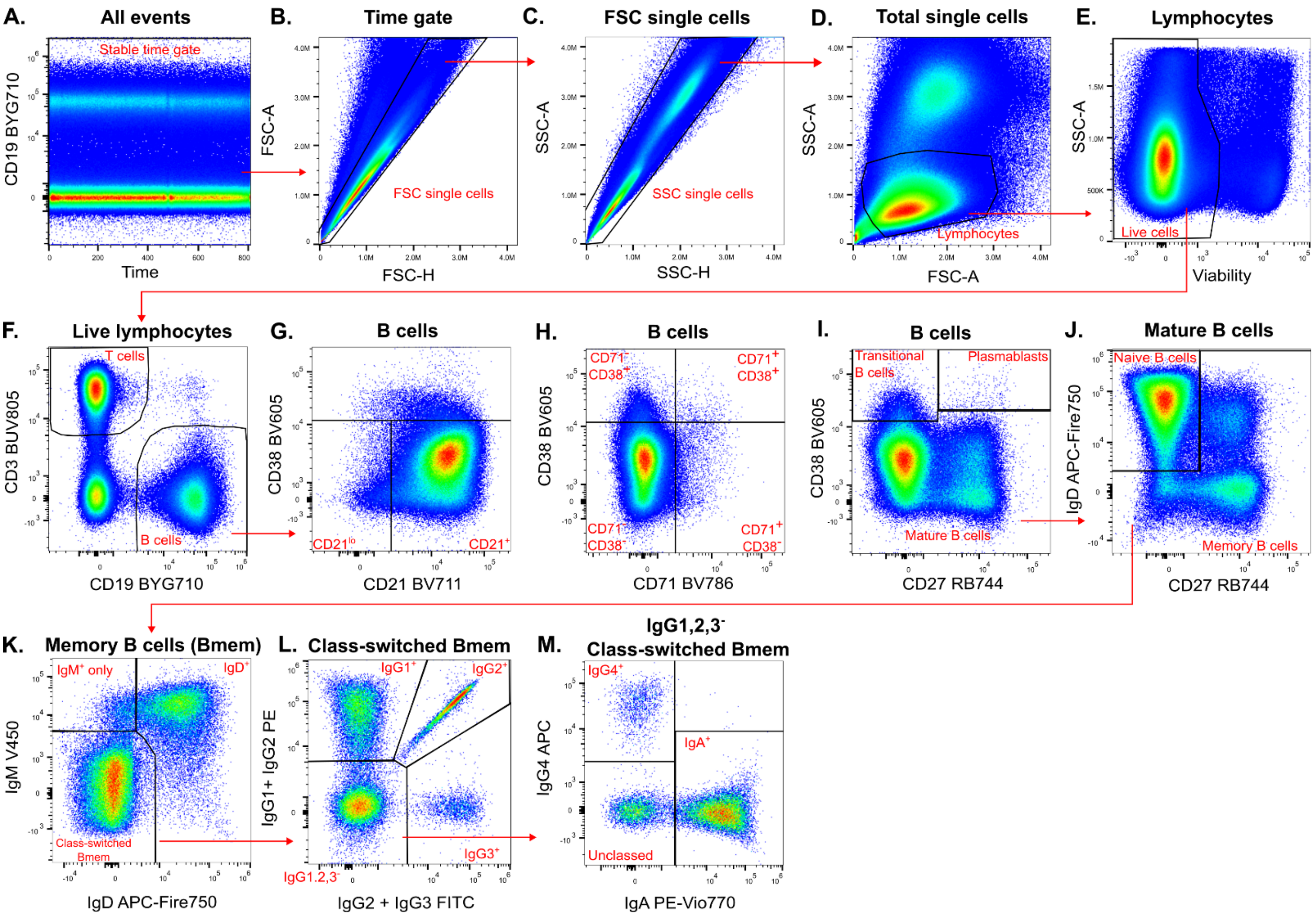


**Supplementary Figure 2. Memory B cell (Bmem) gating strategy**. **(A)** All events were gated on CD19 vs time to ensure a steady signal across acquisition time and to exclude aggregates. **(B-C)** Doublets were excluded on FSC-A vs FSC-H, and subsequently SSC-A vs SSC-H. **(D)** Lymphocytes were gated as $SSC^{lo}FSC^{mid}$. **(E)** Dead cells were excluded on SSC-A vs viability. **(F)** B-cells were gated as CD19+CD3-. **(G)** $CD21^{lo}$ and CD21+ B cells were gated vs CD38. **(H)** CD38 and CD71 expression on B cells was gated in quadrants. **(I)** Transitional and Mature B-cells, and plasmablasts were gated on CD38 vs CD27. **(J)** Naive B-cells and Bmem and were gated using IgD vs CD27 within mature B cells. B mem were additionally phenotyped on CD38, CD21 and CD71 expression using the same gating strategy shown in panels **G** and **H**. **(K)** IgM+IgD+, IgM+ only, and Ig class-switched Bmem were gated on IgM vs IgD. **(L)** IgG1+, IgG2+, IgG3+, and IgG1,2,3- class-switched mature Bmem were gated. **(M)** IgG4+, IgA+, and unclassed Bmem were gated within IgG1,2,3- cells.

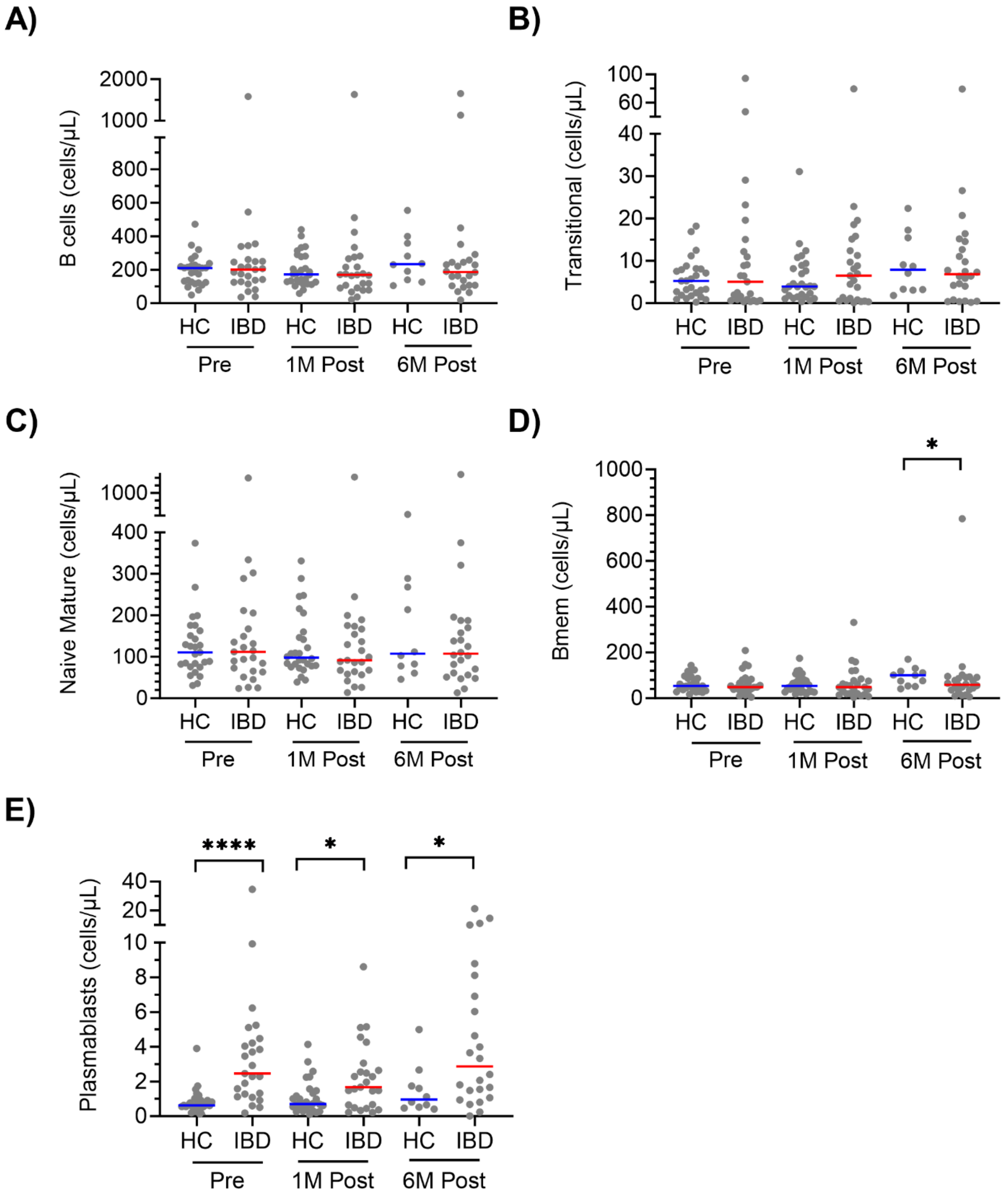


**Supplementary Figure 3. Total B-cell phenotyping before and after vaccination.** Absolute numbers of **(A)** B cells, **(B)** Transitional B cells, **(C)** Naive mature B cells, **(D)** Memory B cells (Bmem). and **(E)** plasmablasts, Horizontal lines depict medians. Statistical significance as calculated by Mann-Whitney test. *p<0.05, ****p<0.0001. HC: Healthy controls. n=27 IBD patients, n=29 HCs (n=11 at 6M post).

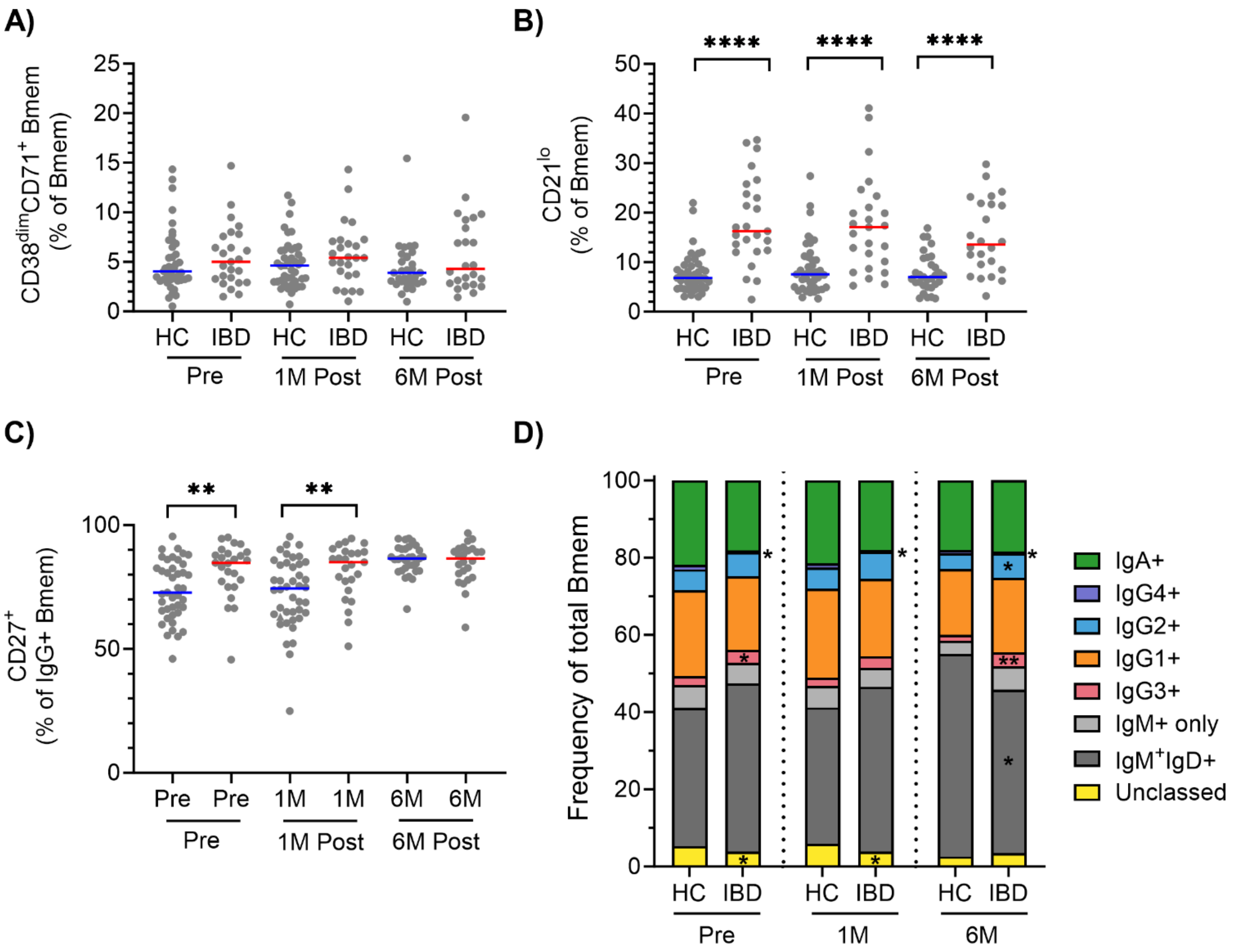


**Supplementary Figure 4. Overall Memory B cell phenotyping before and after vaccination.** Frequencies of; **(A)** recently activated CD21lo Memory B cells (Bmem) expressed as a percentage of total Bmem, **(B)** recently activated $CD38^{dim}CD71^{+}$ Bmem cells expressed as a percentage of total Bmem, and **(C)** secondary germinal center experienced $CD27^{+}$ Bmem expressed as a percentage of $IgG^{+}$ Bmem. **(D)** Frequencies of Ig isotype and IgG subclass expressing subsets within total Bmem.  Data shown as median. Statistical significance as calculated by Mann-Whitney test. *p<0.05, ****p<0.0001

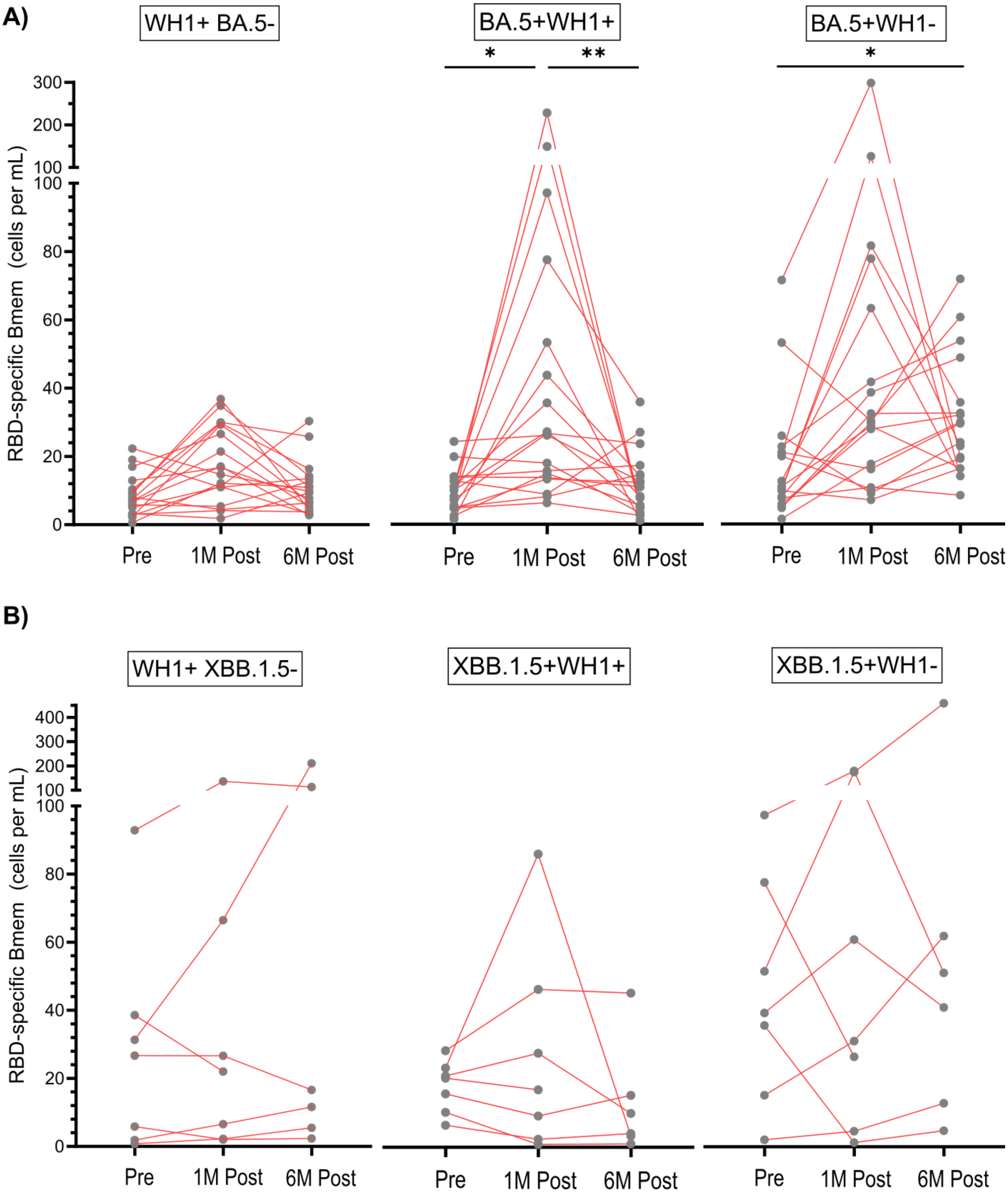


**Supplementary Figure 5. Absolute numbers of RBD-specific Bmem before and after vaccination in IBD patients. (A)** Absolute numbers of WH1+BA.5-, WH1+BA.5+ and WH1-BA.5+ RBD-specific Bmem before and after BA.5 bivalent vaccination. **(B)** Absolute numbers of WH1+XBB.1.5-, WH1+XBB.1.5+ and WH1-XBB.1.5+ RBD-specific Bmem before and after XBB.1.5 monovalent vaccination. Lines connecting paired data. Statistical significance within each subset as calculated by Friedman test with multiple comparisons. *p<0.05, **p<0.01

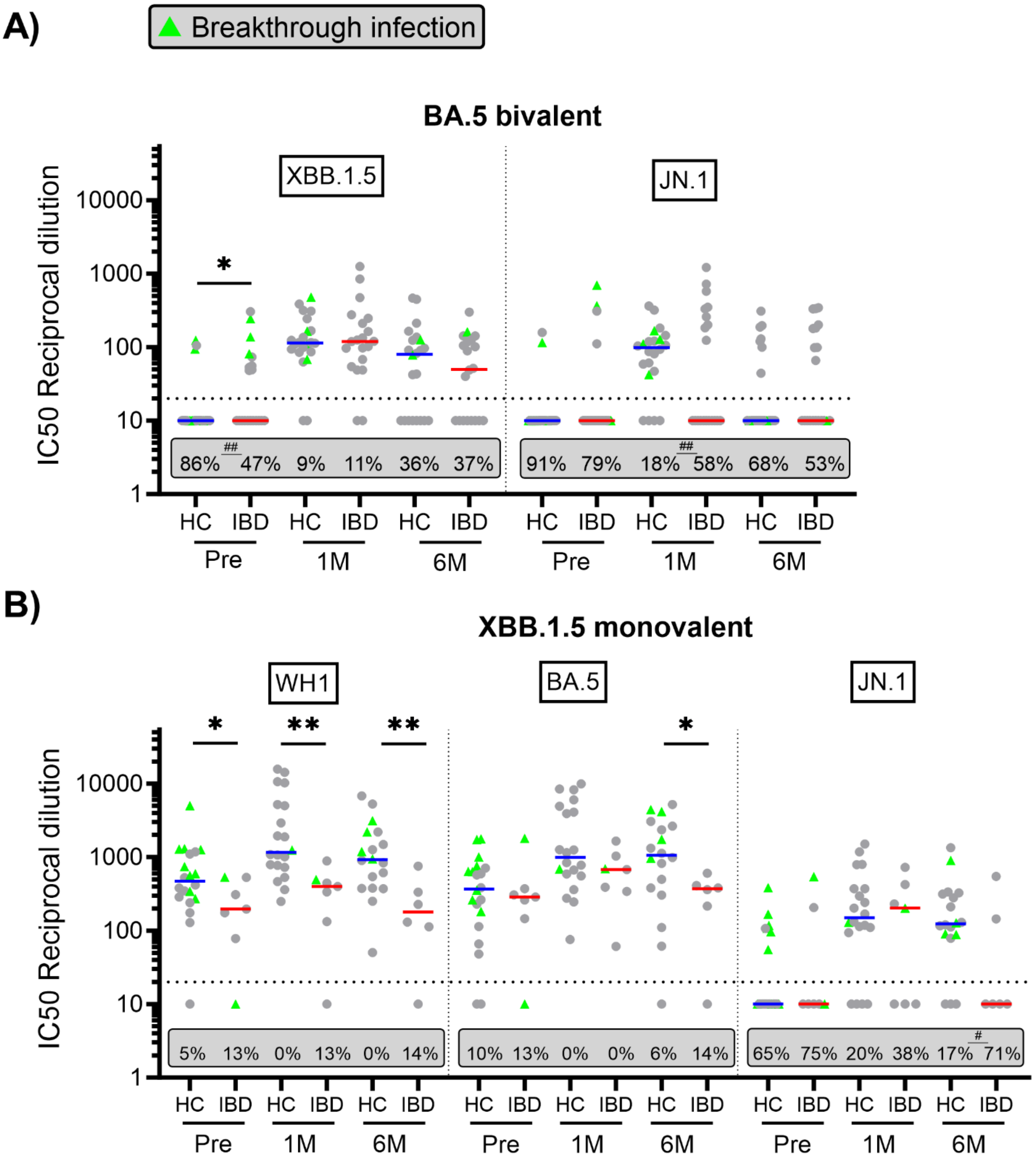


**Supplementary Figure 6. Variant neutralization antibody titers before and after COVID-19 booster vaccination in IBD patients and controls. (A)** Neutralizing antibody titers against XBB.1.5 and JN.1 within individuals who received BA.5 bivalent vaccination (n=19 IBD patients, n=22 healthy controls) **(B)** Neutralizing antibody titers against WH1, BA.5 and JN.1 within individuals who received XBB.1.5 monovalent vaccination (n=7 IBD patients, n=22 healthy controls). Data shown as median. Breakthrough infections were determined as seropositivity to SARS-CoV-2 Nucleoprotein (NCP). Statistical significance between IBD patients and controls as calculated by Mann-Whitney test. ***p<0.001, **p<0.01, *p<0.05. Numbers in grey box represent proportion of individuals below cutoff value of 20 (dotted line) without neutralizing antibodies. Statistical significance between proportions of non-neutralizing responses between patients and controls as calculated by Chi-squared test. #p<0.05, ##p<0.01.

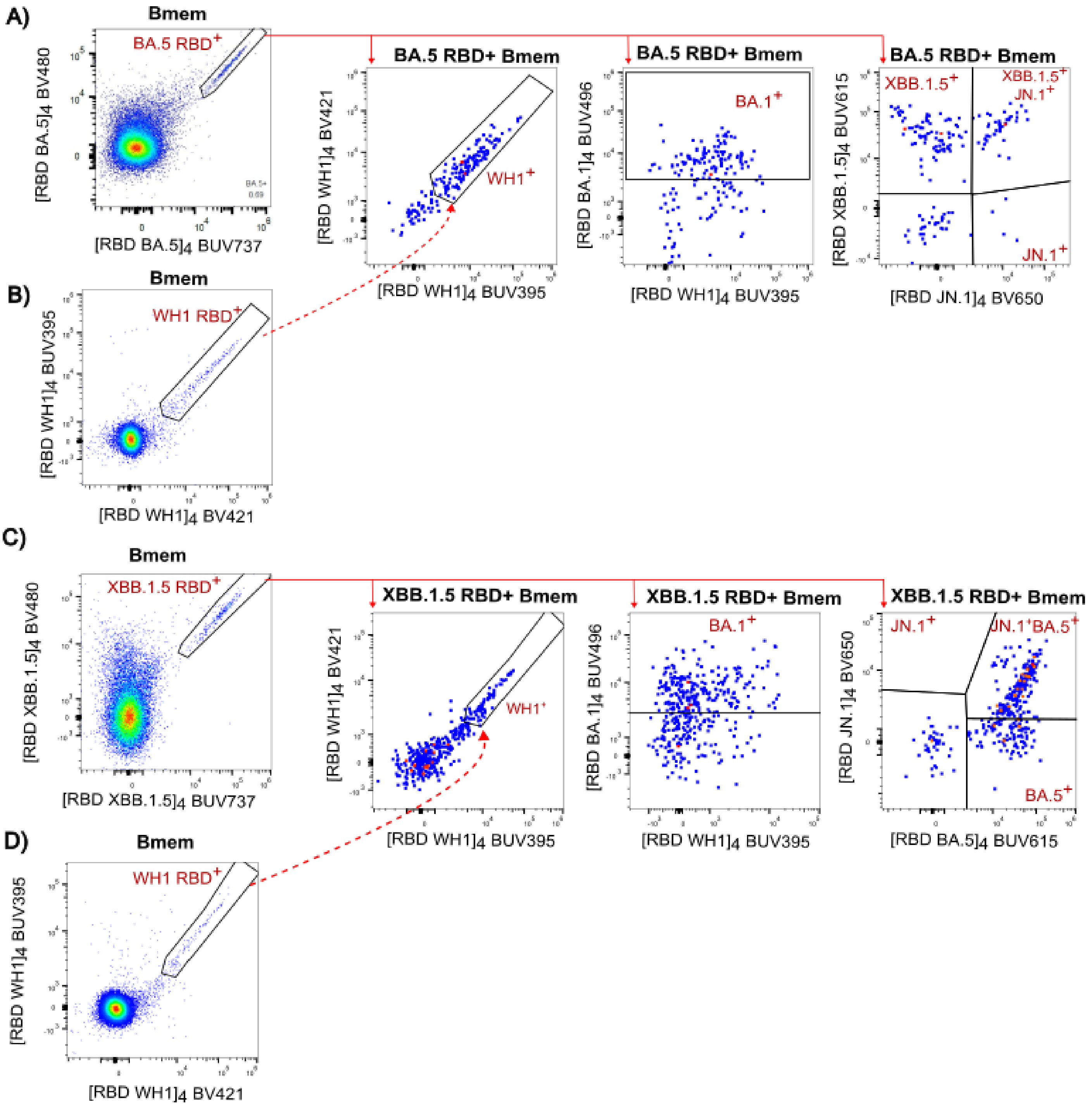


**Supplementary Figure 7. Extended gating strategy for identifying subvariant-binding Bmem. (A)** BA.5 RBD+ Bmem were gated for subvariant binding to WH1, BA.1, XBB.1.5 and JN.1 variants from IBD patients and healthy controls receiving BA.5 bivalent vaccination. **(B)** WH1+ events were gated within Bmem, this gate was also applied to BA.5 RBD+ Bmem to identify WH1 subvariant binding. **(C)** XBB.1.5 RBD+ Bmem were gated for subvariant binding to WH1, BA.1, BA.5 and JN.1 variants from IBD patients and healthy controls receiving XBB.1.5 monovalent vaccination. **(D)** WH1+ events were gated within Bmem, this gate was also applied to XBB.1.5 RBD+ Bmem to identify WH1 subvariant binding.
